# Trends in self-management research in spinal cord injury: A scoping review of study designs and findings

**DOI:** 10.1101/2025.01.06.25320049

**Authors:** Enxhi Qama, Sara Rubinelli, Nicola Diviani

## Abstract

**Context:** Self-management (SM) is essential for people living with a spinal cord injury (SCI) to maintain independence and well-being. While interventions have traditionally focused on medical management, research increasingly recognizes psychosocial and contextual factors. However, how SCI SM research has evolved and whether it aligns with patient-centered approaches remains unclear.

**Objective:** To analyze trends in SCI SM research regarding study designs, settings, populations, and topics, and to synthesize key findings to identify critical gaps.

**Methods:** A scoping review following Arksey and O’Malley’s framework was conducted. PubMed, Science Direct, CINAHL, Scopus, and Cochrane Library were searched for peer-reviewed studies that addressed SCI SM. Study characteristics and trends were quantitatively summarized, while findings were synthesized thematically.

**Results:** Fifty-two studies were included, mostly from the last six years. Research remains predominantly community-based, however there has been a rise in inpatient studies on SM skill-building. The past decade has seen a shift toward qualitative and mixed-methods research, alongside an expansion in topics beyond medical concerns to include emotional well-being. Thematic synthesis revealed three major dimensions shaping SCI SM: *individual factors* (knowledge, psychological well-being, SM integration*), interpersonal and societal influences* (patient-provider relationships, societal contexts), and *technological integration* (enhancement of SM outcomes, usability, and feasibility).

**Conclusions:** This review shows increased and diversified research on SCI SM. Findings emphasize the importance of operationalizing knowledge through skill development, integrating SM into daily routines, and fostering effective communication between patients, caregivers, and healthcare providers. Longitudinal studies from rehabilitation to community are needed to understand and monitor SM over time.

## 1. Introduction

Navigating life after a spinal cord injury (SCI) presents critical challenges, requiring individuals to adopt self-management (SM) strategies to maintain independence and well-being [1–5]. Self-management interventions have been developed to support individuals in this process, providing structured guidance on topics such as skin care [6, 7], medication adherence [8], and physical activity [9, 10]. Literature on these interventions consistently highlight a critical limitation: they primarily focus on medical and symptom management, while neglecting key aspects of real-world self-management, such as emotional management, decision-making, social participation, and adaptation to daily life [11, 12]. Indeed, SM research itself has not adequately addressed these aspects [13–16]. Existing reviews of SM interventions primarily focus on evaluating effectiveness, feasibility, and content—assessing whether interventions improve clinical outcomes, what elements they include, and how they function in different settings [6, 8, 11, 12]. However, they often overlook important findings about how SM is implemented, how participants experience interventions, and the broader contextual factors that shape engagement [11, 12]. Over the past decade, there has been a shift in healthcare research from a biomedical model toward a social model of care, emphasizing patient-centered approaches, health equity, and disability justice [17–19]. Therefore, these elements are crucial for designing interventions that are not only effective in controlled settings but also sustainable and meaningful in everyday life [20, 21].

Beyond the limitations of intervention research, another critical issue is that the way SM is studied influences what we know about it [22, 23]. The choice of study design, outcomes measured, and target populations shape the type of knowledge generated and determine whether research findings translate effectively into practice [24]. Prior reviews have not examined whether research methodologies have evolved to better capture the full complexity of SCI SM. Without understanding research trends, we cannot determine whether SCI SM research is progressing toward a more comprehensive, patient-centered approach or whether it continues to reinforce the same limitations [25, 26].

To address these gaps, this review integrates two complementary objectives: 1) analyzing research trends to examine how SCI SM has been studied over time; and 2) synthesizing findings across all study types to provide a comprehensive understanding of what is known about SCI SM and identify critical knowledge gaps. Doing so, this review offers a more complete picture of how SCI SM is conceptualized, studied, and applied. Examining research trends allows us to assess whether study designs and priorities align with patient needs [27, 28], while synthesizing study findings enables us to capture the full scope of SM beyond intervention effectiveness. Ultimately, this review strives to provide foundational knowledge to advance SCI SM research and ensure that future interventions better reflect the realities of SM in everyday life [29].

## 2. Methods

A scoping review following the five methodological steps suggested by Arksey and O’Malley was conducted [30]: 1) identifying the research questions, 2) identifying relevant studies, 3) study selection, 4) charting the data and 5) collecting, summarizing, and reporting the results. Arksey and O’Malley methodology provides a structured and systematic approach for conducting a scoping review, allowing for an extensive overview of the current state of research on a given topic [30]. Furthermore, the Preferred Reporting Items for Systematic Reviews and Meta-Analysis Extension for Scoping Reviews (PRISMA-ScR) checklist for reporting the steps was adopted [31] (see Supplementary table, PRISMA-ScR Checklist).

### 2.1 Research question

For this review our research questions are as follows:

1. How has SCI SM research evolved over time in terms of study designs, settings, target populations, and commonly studied topics, and to what extent does this evolution reflect a shift toward social models of care and patient-centered approaches?
2. What are the key themes and insights emerging from SCI SM research, particularly in relation to how individuals experience SM into daily life and navigate contextual and structural barriers?

### 2.2 Search for relevant studies

Relevant databases included Pubmed, Science Direct, CINAHL, Scopus and the Cochrane Library were searched on May 2024. The search terms included keywords related to SCI and SM. Boolean operators such as “OR” and “AND” were used to combine the search terms (see Table 1). We purposively focused our search on studies that explicitly mentioned ‘self-management’ or its synonyms. As a result, studies that did not use these exact terms, even if they discussed related concepts or components of SM, were not retrieved. By narrowing our search, we ensured that our analysis was conceptually focused on SM as a distinct construct.

A full search was conducted to retrieve all relevant articles and based the selection process on the following eligibility criteria: (1) all study designs, to ensure a comprehensive understanding of research trends and findings, (2) studies in English, (3) studies published in the last 21 years, reflecting the emergence and development of SM research in SCI, as identified from a preliminary search (4) with adult participants, as these differ significantly from pediatric contexts (5) only original, peer-reviewed studies to ensure scientific rigor of the evidence base (excluding protocols); and (6) the studies needed to explicitly address SM in SCI, either as primary focus or as part of their findings, to align with the study’s objective.

### 2.3 Selecting the relevant studies

A reference manager was used to export the identified studies and eliminate duplicate records. Following this, titles and abstracts were screened by two reviewers independently to determine study eligibility based on predefined criteria. In cases of disagreement, a third reviewer was consulted to reach consensus and ensure consistency. Then the full text of a random sample of 10 studies was independently conducted by two reviewers. This step aimed to evaluate inter-reviewer agreement and refine the application of eligibility criteria, ensuring clarity and consistency before proceeding to full-text screening phase. Any discrepancies in this step were resolved through discussion and consensus, with input from a third reviewer if necessary.

Finally, a full text screening was done by one reviewer of all remaining studies against the eligibility criteria (see Figure 1. PRISMA flow chart).

### 2.4 Charting the data

A standardized table was developed by two reviewers to extract relevant information from the eligible articles. Each reviewer independently extracted data from all articles, and discrepancies were identified and resolved through a consensus-based approach to ensure accuracy and consistency. The extracted data were then organized into two separate tables. The first table details the formal characteristics of the studies, including author, country, year, study design, population, setting, health topic addressed, data collection instruments, outcomes, and predictive measures. The second table summarizes the main results of each study, along with the SM tasks explored, categorized according to Lorig et al.’s framework of SM in chronic conditions [29], as identified in the study’s stated focus (see Table 2 and Table 3). The Lorig framework is one of the most widely used conceptual models in SM research, originally developed to classify the key domains of SM across chronic conditions. It categorizes SM into three primary domains: 1) Medical management, including tasks that involve direct physiological management of a condition, such as medication adherence, symptom monitoring, physical therapy, and pressure ulcer prevention. 2) Role management, including the ability to adapt and maintain meaningful life roles despite the condition, including returning to work, engaging in social and family life, and managing household responsibilities. 3) Emotional management, including psychological and emotional coping strategies, such as adjusting to a new identity post-injury, dealing with stress, anxiety, or depression, and building resilience [32].

This framework is widely used in research on SM interventions because it provides a structured way to assess how individuals manage their condition beyond medical tasks alone [33]. Prior research in diabetes, arthritis, and other chronic conditions has successfully used this framework to design and evaluate SM programs [34, 35]. To systematically categorize SM tasks within this framework, we assessed each study’s stated objectives, reported findings, and descriptions of SM activities. Studies were mapped into one or more categories if they explicitly addressed the domains, either as an intervention focus or as part of participants’ reported experiences.

Additionally, we acknowledge that some included studies refer to the same intervention [36–44] or are studies that share the same dataset with overlapping participants [45–50]. To ensure clarity and avoid duplication in our synthesis, for trends analysis, these studies were counted only once when summarizing settings, and topics. For thematic synthesis of findings, each study were analyzed separately if they provided unique findings (e.g., different aspects of SM, patient experiences, or intervention effectiveness).

### 2.5 Collating summarizing, and reporting the results

For the first research question, we systematically examined how research in SCI SM has evolved over time by categorizing studies based on their designs, settings, target populations, and health topics. The frequencies of different study characteristics were quantified, and temporal trends were assessed by analyzing publication patterns over the years. We identified shifts in research priorities, study methodologies, and populations studied, highlighting whether newer research aligns with contemporary models of patient-centered care and SM.

For the second research question (main findings), we extracted data on self-management experiences, intervention effectiveness, challenges in implementation, and contextual factors influencing SM behaviors. To ensure consistency in the thematic analysis, the results of each study were first summarized by one reviewer in a structured format, and these summaries were used as the data source for coding and thematic synthesis. This ensured that extracted findings were systematically categorized and synthesized based on shared meanings across studies. The summarized results data from the studies’ results were analyzed using a mixed-methods approach. This approach focuses on creating a body of empirical research based on the findings of both primary qualitative and quantitative studies, which makes it appropriate to explore what is known about a specific phenomenon and direct future research and practice [51–53].

Specifically, an integrated design was used, which aims to make results from different methodologies comparable by means of transformation of the data, which in our case is “qualitizing” quantitative results and making a narrative summary of the combined results as suggested by Sandelowski et. al [51]. For studies with quantitative methods, the authors’ stated interpretations of their numeric findings were extracted rather than conducting new analyses or reinterpreting the data (see Table 3). After qualitization, all data were coded inductively to identify overarching themes. The extracted data was independently coded by two reviewers, using an inductive approach to identify subthemes and themes. Discrepancies between the two coders were resolved through discussion and consensus, ensuring consistency in thematic identification. When disagreements persisted, a third team member was consulted for final adjudication. Finally, the codes were grouped into themes based on shared meanings or ideas from diverse study designs and synthesized into a narrative (see Figure 2 for an example of theme development).

## 3. Results

### 3.1 Study selection

After screening the abstracts of 2792 articles, 92 were included for full-text screening. Of these, 41 studies were deemed ineligible. Ultimately, 52 articles in total were included for synthesis, as illustrated in Figure 1. It is worth mentioning that several studies included in our review refer to the same intervention [36–44], and some refer to studies that have the same data collection with the same participants [45–50].

The studies were conducted across various countries, with the majority (n=18) in Canada [43–50, 54–63], followed by the United States (n=17) [38–42, 64–75]. Other countries included Switzerland (n=5) [36, 37, 76–78], the Netherlands (n=3) [79–81], Thailand (n=2) [82, 83], China (n=2) [84, 85], and one each in France [86], South Africa [87], Australia [88], Bangladesh [89], and the UK [90]. Despite the international growth of SM research, we still see a lower representation from low- and middle-income countries.

The included studies covered a 21-year period ranging from 2003-2024, with the large majority (n=34) conducted in the last six years [28–34, 37–41, 43, 45, 47, 48, 51–53, 57, 60–62, 65, 66, 68, 69, 71–76, 79] (see Figure 3). While this recent increase may suggest a growing recognition of the importance of SM research in SCI, there remains an imbalance in focus areas and study approaches, which we explore below.

### 3.2 Study characteristics and trends in research on SM in SCI

#### 3.2.1 Shifts in study designs

Over the past two decades, SCI SM research has evolved to include a broader range of study designs. While early studies primarily relied on RCTs, recent research has increasingly adopted qualitative and mixed-methods approaches to capture patient experiences and contextual factors affecting SM (see Figure 4). Qualitative studies (n=23, 44%) were the most common and have grown in prevalence over the past five years (n=18). These studies explored SM strategies, implementation challenges, lived experiences, and intervention feasibility [37, 40, 45–50, 54, 55, 59–61, 64, 70, 73, 74, 77, 78, 81, 87–89]. Feasibility studies (n=10, 19%) emerged as the second-largest category, with seven conducted in the past five years. These studies used quantitative (n=5) and mixed-methods (n=5) designs [42, 43, 56, 62, 63, 65, 68, 69, 71, 72].

RCTs (n=8, 15%), while essential for evaluating intervention effectiveness, were fewer in number [38, 39, 64, 66, 67, 75, 80, 84]. Most were conducted between 2012 and 2017, with only three in the last five years. The relatively low number of RCTs (n=8) compared to the growing number of feasibility studies (seven in the last five years) may suggest a research focus on refining SM interventions, assessing usability, and improving implementation strategies before conducting large-scale effectiveness trials.

Observational cross-sectional studies (n=6, 11%) examined patient-reported SM strategies, prevalence of depression, and patient activation measures, often providing valuable but static snapshots of SM behaviors [58, 79, 82, 83, 85, 90].

Intervention development studies (n=5, 10%) explored the design and refinement of SM programs before implementation. These studies often focused on program components, user experiences, and system integration [36, 41, 44, 57, 76].

#### 3.2.2 Changes in study settings

Most studies were community-based (n=35, 81%), reflecting the long-standing emphasis on post-discharge SM. However, inpatient-focused studies have increased in recent years (n=5, 10%), signaling recognition of the importance of early-phase SM skill-building within rehabilitation settings. Community-based studies (n=35) included a mix of intervention and observational designs, covering various SM topics such as medication adherence and pressure ulcer prevention [36–46, 49, 50, 54, 58–61, 63, 66–77, 79–82, 84–90]. Inpatient studies (n=5) explored early-stage SM training and structured intervention implementation in rehabilitation settings [55, 57, 62, 64, 83].

Mixed-setting studies (n=3, 6%) examined how SM skills developed in inpatient settings translated into home and community environments [56, 65, 78]. Outpatient studies (n=1, 4%) focused on continued follow-up care and specialized SM programs [47, 48]. The recent increase in inpatient studies suggests a shift toward early-stage SM interventions, rather than relying solely on post-discharge community-based support.

#### 3.2.3 Changes in research population

While patients with SCI remain the primary study population (n=2260 individuals across studies), recent research incorporates caregivers (n=67), healthcare professionals (n=143), and other stakeholders (n=82, including experts, researchers, and policymakers). This diversification may indicate a growing recognition that SM is influenced not only by individual behaviors but also by social and structural factors [13]. Studies involving caregivers and healthcare providers focus on their role in supporting patient SM, training needs, and system-level barriers to effective SM implementation [47, 48, 65, 78]

#### 3.2.4 Shifts in research focus

SCI SM research has traditionally emphasized medical management, particularly pressure injuries, bladder and bowel care, and medication adherence. However, recent studies highlight psychological and social aspects of SM, aligning with broader shifts in disability research. Pressure injuries (n=13, 27%) remain the most frequently studied topic [36, 37, 61, 64, 70, 71, 76–79, 81, 85, 86, 90]. Bladder and bowel management (n=5, 13%) has gained attention, particularly in recent intervention research [40–42, 67, 83, 88]. Medication adherence (n=2, 8%) remains a key focus, though often integrated into broader SM programs [45–48]. Mental health (n=2, 4%) has only recently emerged as a research priority, despite the well-documented psychological burden of SCI [82, 84]. General SM approaches (n=18, 40%) include studies examining holistic SM, patient activation, and social participation [38, 39, 43, 44, 49, 50, 54–59, 62, 65, 68, 69, 72, 74, 75, 87, 89].

#### 3.2.5 Gaps in SCI SM research

Despite the broadening of research methodologies and focus areas, significant gaps remain. First, depression and mental health remain underrepresented, despite being critical for long-term SCI management. Second, while Lorig’s framework classifies SM into medical, role, and emotional management, most studies focus only on medical tasks. Only one study specifically explored the role management, indicating a lack of research on how people integrate SCI management into daily roles and relationships [89]. Third, research primarily assesses short-term intervention outcomes, with no focus on long-term maintenance of SM behaviors. Fourth, despite increasing recognition of the role of caregivers and healthcare professionals in SCI SM, limited research explores how to train and support them effectively.

### 3.3 Narrative synthesis of studies’ findings

From the thematic analysis, three themes emerged capturing the most significant insights from SCI self-management research: Individual factors; Interpersonal and social influences; and Technological integration. *Individual factors* included knowledge, psychological well-being, and SM integration in daily life. *Interpersonal and societal influences* examined external social and relational aspects affecting SM in individuals with SCI. *Technological integration* explained the potential of apps and programs to improve SCI health outcomes and elements affecting their uptake and use (see Table 4). It is worth noting that the studies cited in each thematic category serve as illustrative examples rather than an exhaustive list of all relevant studies.

#### 3.3.1.1 Individual factors

This theme explored three personal elements influencing SM in individuals with SCI. It included 1) knowledge in the form of educational information and applied knowledge, 2) the impact of psychological well-being on SM practices, and the 3) integration of SM into one’s daily life. Each element is described in detail below.

##### 3.3.1.1.1 Knowledge at the cornerstone of SM

Knowledge was pivotal in the SM of SCI, as highlighted by several studies, revealing its importance across various levels: individual, caregiver, and health professional [36, 37, 39–41, 44, 45, 47–50, 54, 55, 57–61, 63, 66, 73, 74, 76, 77, 79, 81, 85, 86, 88]. More concretely, two facets of knowledge stood out: educational information and applied knowledge.

Educational materials often covered complications [61, 76] risk factors [77, 86], medication management [47, 48] and preventive measures like catheterization and physical activity [40, 60]. An important consideration was the health literacy levels of individuals using this information [57] and the necessity of avoiding medical jargon to ensure comprehension [59, 73]. The source of information was also critical, emphasizing the need for platforms to offer credible resources [39] and filter information appropriately [44]. For instance, peers and health professionals [39, 73] played a pivotal role in knowledge sharing, particularly in confidently delivering information [55, 74, 81], which, as showed by Cadel et al., could impact adherence to medication management [45, 48].

Applied knowledge involved understanding one’s body and recognizing sensations, such as in the case of for intermittent urinary catheter users [40] and the early detection of pressure ulcers [86]. Studies emphasized action-planning, goal setting, information seeking, problem-solving and decision-making skills [45, 54, 58, 60, 63, 74, 86] as essential for more effective SM in SCI. Applied knowledge also extended to caregivers, encompassing skill sets for prevention, monitoring, and managing complications [36, 50, 73]. For instance, Pryor et al. showed that the support provided by caregivers was pivotal for integrating practices like bowel care, advocating for training caregivers in collaborative problem-solving initiatives to enhance SM [88].

##### 3.3.1.1.2 Psychological well-being as an important element in SM

The significance of psychological health emerged as a crucial aspect, underscoring its impact on individuals’ ability to effectively manage SCI [49, 74, 81, 86, 87, 89].

Feelings of anxiety, negative mood, frustration, and depression were commonly reported by individuals with SCI and were recognized as serious barriers to SM [49, 86]. Pilusa et al. explored the experiences of individuals with SCI in managing secondary conditions, showing feelings of frustration and hopelessness when SM strategies proved ineffective [87]. Similarly, Van Gaal et al. found that anxiety and frustration often arose from strained relationships with caregivers during efforts to prevent and manage pressure ulcer [81].

Peers and caregivers served as vital sources of motivation, encouragement, and advocacy for individuals with SCI, helping to mitigate mental health challenges [49, 74, 81]. Begum et al. identified engaging in recreational activities, maintaining a sense of identity and purpose, and receiving societal rewards such as respect and recognition for contribution in social environments, as effective means of managing emotions in individuals with SCI [89].

##### 3.3.1.1.3 Self-management of SCI as part of daily life

A persistent element in the included studies was the integration of SM activities into daily life, which fostered confidence, skills, and lifestyle adjustments [40, 45–48, 50, 54, 58–60, 68, 86, 88, 89]. Individuals with SCI navigated several SM tasks [50, 59, 89], including catheterization [40], medication management [45–48], pressure ulcers [86], bowl care [88], all requiring constant monitoring and adjustment to fit into daily life. For instance, Cadel et al. highlighted participants’ adeptness at problem-solving and decision-making skills, enabling them to tailor medication management routines to their daily lives by utilizing strategies such as organization and planning to ensure adherence [46]. Flexibility in scheduling was crucial, allowing individuals to accommodate other responsibilities like work or family time [45, 86].

#### 3.3.1.2 Interpersonal and societal influences

This theme examined the external social and relational factors that affect SM in individuals with SCI. It showed the crucial role of trust and communication between patients and healthcare providers in facilitating effective SM. Furthermore, it explored the influence of broader societal dynamics and attitudes on SM practices, illustrating how they could impact health outcomes.

##### 3.3.1.2.1 Patient-provider relationship influences SM

The relationship and communication between patients and healthcare professionals emerged as crucial factors in the SM of SCI, as emphasized by several studies in our review [47, 48, 77, 78].

Studies focusing on medication and pressure ulcer management underscored the significance of trust, open communication, and collaborative decision-making between patients and providers. For instance, Zanini et al. explored how different collaboration styles between patients and providers corresponded to different patient approaches to their issues, such as being thoughtful, selective, or delegative [77]. Furthermore, healthcare professionals emphasized the importance of defining responsibilities, negotiating priorities, setting goals, and cultivating mutual trust and respect to build effective partnerships with individuals with SCI [78].

##### 3.3.1.2.2 Societal context shapes SM of SCI

The societal context played a significant role in shaping the landscape of SM in SCI, as reflected in various study findings [40, 45, 57, 59, 81, 86, 89].

For instance, users of intermittent urinary catheter faced challenges in adapting to social situations, particularly in intimate relationships, and experience associated feelings of embarrassment [40]. Gourlan et al. highlighted the social ramifications of pressure ulcers, including withdrawal and isolation, which significantly impeded daily activities [86]. Similarly, participants at Van Gaal et al.’s study noted that managing pressure ulcers often required balancing family life, work and hobbies, as many preventive measures were difficult to apply in certain circumstances [81].

Begum et al. proposed an additional aspect in their SM model, named “management of social complexities”, which included relocation to new environments and considering disability-friendly settings free of discrimination and negative attitudes. They advocated for efforts to promote accessibility and defend rights to enable greater participation in daily life [89]. This focus was especially crucial given the evidence of social isolation post-discharge [57].

#### 3.3.1.3 Technological integration

This theme investigated the role of technology in enhancing SM for individuals with SCI. It assessed how the effectiveness of SM apps and programs could significantly improve health outcomes by providing real-time monitoring and personalized feedback. Besides, it delved into the usability and feasibility elements that influence the adoption and sustained use of these digital tools, ensuring they are accessible and practical for users.

##### 3.3.1.3.1 Apps and programs enhance SM outcomes

The effectiveness of apps and programs in improving many outcomes in SM in SCI was a common topic throughout our studies [38, 42, 43, 56, 67, 75, 84]. These digital tools have been shown to reduce the risk of secondary complications [38, 67, 84], and foster positive changes in self-efficacy, resilience, emotional status, and health literacy [43, 75]. Another explored outcome was the level of confidence in performing a SM activity. For instance, Wilde et al. demonstrated that a web-based intermittent catheter SM intervention increased awareness of fluid intake and enhanced confidence [42]. Similarly, MacGillivray et al., observed improved confidence in bowel management over time through the use of their mobile app [56].

##### 3.3.1.3.2 Usability and feasibility elements influence the adoption of apps and programs

This last group of studies showed how the successful adoption of apps and programs for SM in SCI depended heavily on their usability and feasibility [36, 42, 43, 61, 62, 65, 69–72, 83].

High usability scores have been frequently highlighted, with studies emphasizing the importance of interesting interfaces [36, 69, 72, 83], intuitive and straightforward navigation [62, 71]. Participants reported increased engagement and improved confidence when using apps that are easy to navigate and visually appealing [65]. Key features contributing to ease of use included independence from internet connectivity, ensuring accessibility in various settings [62], appropriate font sizes for readability [69, 72], and well-designed button arrangements and page navigation for intuitive use [72].

Regarding feasibility, most studies reported high retention and adherence rates [43, 70, 71, 83]. Successful programs, like the one studied by Wilde et al., incorporated features such as diaries, educational materials, calls, and interaction with nurses, which were easily adopted and valued by participants [42]. However, challenges to feasibility included connectivity issues [71], and misuse of apps by participants, such as contacting health professionals for social interaction or irrelevant information.

Furthermore, maintenance and financial concerns were identified as potential barriers to feasibility as showed by Amann et al. [36].

## 4. Discussion

In this scoping review, we analyzed the evolution of research designs, study trends, and key findings in SCI SM. Our results indicate a shift toward more varied methodologies, with a notable increase in qualitative and mixed-methods studies alongside feasibility studies assessing intervention usability. While RCTs remain underrepresented, feasibility studies have increased, suggesting an effort to assess usability before large-scale trials. Additionally, earlier research predominantly focused on post-discharge, community-based SM. More recent studies incorporate SM training during inpatient rehabilitation, acknowledging the importance of early-phase skill development. Moreover, medical management remains the dominant focus, with continued emphasis on pressure injuries, bladder and bowel care, and medication adherence. However, mental health concerns like depression are emerging, reflecting a growing understanding of the emotional burden of SCI. Social and role management remain underexplored, despite their critical role in long-term SM success.

From an implementation science perspective, this review contributes to knowledge synthesis, serving as a first step towards future intervention implementation [91, 92]. First, while knowledge and information remain central to SM research [6–8, 11, 16], our review emphasizes the need to operationalize this knowledge through skills like action planning, information seeking, problem-solving and decision-making, which are fundamental to effective SM [29, 93–96]. For instance, in heart failure research, decision-making is viewed as a naturalistic process, deeply embedded in personal contexts [97]. Daley et al. describe these processes in a three steps framework of monitoring, interpreting symptoms and acting accordingly [98]. Investigating whether these strategies could be adapted for SCI could offer valuable insights. Secondly, current interventions often treat SM activities as separate from everyday life [8, 11, 12], which is particularly challenging for individuals managing multiple complications or coexisting conditions [99]. This fragmentation forces individuals to compromise personal or social pursuits for SM activities [96, 99–102]. For instance, Gross-Hemmi et al. demonstrated low participation and a decrease of leisure activities over a 5-year time period in individuals with SCI [103]. Thirdly, SM interventions often address issues perceived as problematic by healthcare professionals or institutions, which may not align with what matters for the patients [104–107]. Studies show that patients prefer goal setting related to everyday life, while healthcare providers tend to use the hospital setting as their point of reference [108, 109]. Our review underscores the importance that patients, caregivers and healthcare providers give to the development of communication frameworks that would facilitate better adherence to SM practices. For instance, such interventions could be adapted to fit not only the health needs of patients, but also the specific circumstances of family, job, and social engagements [110–112].

### 4.1 Gaps in the literature and further research

Our review identified important gaps and critical areas for future research.

While our work is descriptive and theoretical, it indirectly supports the initial steps of the implementation process for SM interventions, as outlined in Graham et al.’s Knowledge-to-Action (KTA) framework. This framework presents actionable phases for translating knowledge into practice, such as selecting relevant knowledge, adapting it to the context, tailoring interventions, and monitoring, evaluating and sustaining knowledge use [91].

In terms of relevant knowledge, emotional aspects of SM remain underexplored. Despite their importance [113], only two studies addressed managing depression [82, 84], showing the need for greater emphasis on psychological well-being in SM interventions. While skin management is well-represented, other areas like bowel management, physical activity, problem-solving, decision-making need more attention.

There is also limited attention to skill development for health professionals and caregivers involved in SM [47, 48, 65, 78]. Despite frequent mentions of caregiver burnout and inadequate training [49, 73, 88], few studies directly address the educational needs of these groups. Geographical gaps are also noted, with a predominance of studies from Western countries. More geographically diverse research could uncover how socio-cultural factors, such as spirituality or traditional roles, influence SM practices [114, 115]. Study design limitations also hinder the applicability of findings. RCTs are underrepresented, with only eight identified. These trials often focus on outcomes like quality of life, while neglecting psychosocial and long-term impacts of SM interventions [116].

The KTA framework’s actionable phase of adaptation of knowledge to the local context aligns with our results for the need of individuals to integrate SM into daily life. Future research should explore how individuals use health knowledge in real-world decision-making, accounting for personal, societal, and contextual factors [13, 117].

Additionally stigma and social barriers remain significant challenges requiring targeted mitigation strategies [118]. To advance SM research and implementation a shift toward more social and patient-centered approaches of care is required [17–19]. Community-Based Participatory Research (CBPR) offers a promising strategy, as it actively involves individuals with SCI in designing, implementing, and evaluating SM programs [119, 120]. Integrating CBPR could enhance intervention relevance, improve real-world feasibility, and ensure that SCI SM research reflects patient priorities and lived experiences [121].

Finally, our review suggests that filling the gap in longitudinal research designs could contribute to the KTA’s phase of monitoring and evaluating knowledge. Longitudinal studies are important for understanding how SM practices evolve over time and across different contexts [122]. Additionally, advancements in artificial intelligence and large language models hold high promise for monitoring and evaluating large amounts of date over time; studying their usability and feasibility for this purpose is a critical next step in implementation of these developing technologies into SM interventions [12]. Conversational agents for example could support SM by promoting adherence, providing information and motivation, and even offering reminders or entertainment [123–126].

### 4.2 Strengths and limitations

To our knowledge, this is the first review that includes a wide range of studies with various designs and methodologies, to then report on the current trends of research in SM of SCI and their findings, including individuals with SCI, caregivers, and health professionals as study participants. This review has some limitations. First, we limited our search to certain eligibility criteria, which may have excluded relevant research, potentially affecting the comprehensiveness of our review. Therefore, the breadth of included studies may have resulted in some limitations in depth. For instance, our search strategy focused on studies explicitly mentioning ‘self-management,’ which may have excluded research exploring related concepts under different terminology. Future reviews could explore these nuances in greater detail. Nonetheless, we attempted to mitigate this limitation by conducting our search across multiple databases and including as many studies as possible in our screening process. Second, we did not conduct a quality assessment of the included studies, which could potentially undermine the robustness of our findings. Nevertheless, this study was designed as a scoping review, with the primary goal of providing a general overview of the current state of research on SM in SCI, rather than critically evaluating study strengths and limitations or assessing the effectiveness of specific interventions.

## 5. Conclusion

This scoping review highlights the recent focus of research on designing and developing SM interventions, the growing use of qualitative and mixed-methods approaches, and the increasing attention to inpatient settings and mental health topics like depression. Key findings emphasize the importance of operationalizing knowledge through skill development, integrating SM into daily routines, and fostering effective communication between patients, caregivers, and healthcare providers. Additionally, significant gaps remain in areas such as the emotional dimensions of SM and the educational needs of caregivers and health professionals. By expanding research in these areas, we can better tailor SM interventions to meet the complex needs of individuals with SCI.

## Data Availability

All data produced in the present work are contained in the manuscript

