## Supplemental Table 1 for "Trends in self-management research in spinal cord injury: A scoping review of study designs and findings"

| **Database** | **Search string combination** |
| --- | --- |
| PubMed | ("self management" OR "self care") AND ("spinal cord injury"[Title/Abstract] OR SCI[Title/Abstract] OR paraplegia[Title/Abstract] OR tetraplegia[Title/Abstract] OR quadriplegia[Title/Abstract]) |
| Cinahl | TX ( "self management" OR "self care" ) AND AB ( "spinal cord injury" OR SCI OR paraplegia OR tetraplegia OR quadriplegia ) |
| Scopus | ALL ( "self management" OR "self care" ) AND TITLE-ABS-KEY ( "spinal cord injury" OR sci OR paraplegia OR tetraplegia OR quadriplegia ) |
| Science Direct | Title, abstract, keywords: ("self management" OR "self-management" OR "self-care" OR "self care") AND ("spinal cord injury" OR SCI OR paraplegia OR tetraplegia OR quadriplegia) |
| Cochrane | ("self management" OR "self care") AND ("spinal cord injury" OR SCI OR paraplegia OR tetraplegia OR quadriplegia):ti,ab,kw |
