## Supplemental Table 2 for "Trends in self-management research in spinal cord injury: A scoping review of study designs and findings"

| Study ID | Type of study | Author, Year | Country | Setting | Year | Type of article | Research design | Sample | Health topic | Data collection/instrument | Outcomes: | Predictor measures/confounders |
| --- | --- | --- | --- | --- | --- | --- | --- | --- | --- | --- | --- | --- |
| S1 | Mixed-methods | Allin et al. [43][44] | Canada | Community | 2020 | Mixed-methods | Feasibility study | Patients (n=11) | Non-specific | Questionnaire | - Numeric outcome:   - Retention and adherence rates - Narrative outcome:   - Program impact and role of health coaches   - Feasibility measures, impact measures | n/a |
|  |  |  |  |  | 2018 | Qualitative | Intervention development | Patients (n=9), researchers (n=7) | Non-specific | Meetings discussions | - Numeric outcome: n/a - Narrative outcome:   - Stage One - Exploration: Self-Management and internet; Resource review responses   - Stage Two - Discovery: A community-curated resource database; Online information navigators; Online groups   - Stage Three - Prototyping: Stars and up-voting; Expertise points and activity feeds | n/a |
| S2 | Mixed-methods | Amann et al. [36][37] | Switzerland | Community | 2020 | Mixed-methods | Intervention development | Project team, Parahelp, home care provider, wound specialist, nutritionist, user experience designer, design sprints (n=19) Patients (n=15) | Pressure injury | Questionnaire and interviews | - Numeric outcome:   - Usability rates - Narrative outcome:   - App design and features   - Usability assessment   - Prototype features   - Feedback and user ratings   - Accessibility and user experience | n/a |
|  |  |  |  |  | 2020 | Qualitative | Qualitative design | Patients (n=15), health professionals (n=13) | Pressure injury | Interview | - Numeric outcome: n/a - Narrative outcome:   - Benefits promoting uptake: A companion for newly injured individuals, An emergency kit and motivational support, A guide for informal caregivers and family members   - Challenges impeding adoption: Motivating individuals to use the app, Concerns about misuse and abuse of the app, Organizational and maintenance challenges | n/a |
| S3 | Quantitative | Baniya et al. [82]  2022 | Thailand | Community |  | Quantitative | Observational design cross-sectional | Patients (n=115) | Depression | Questionnaire | - Numeric outcome:   - Prevalence and severity of depressive mood   - Self-management methods to cope with their depressive mood - Narrative outcome: n/a | n/a |
| S4 | Qualitative | Begum et al. [89]  2022 | Bangladesh | Community |  | Qualitative | Qualitative design | Patients (n=14) caregivers (n=14) | Non-specific | Photoelicitation discussions | - Numeric outcome: n/a - Narrative outcome:   - Medical Management: Developing and following consistent routines and habits, Using health-promoting properties of daily activities   - Emotional Management: Developing authority over feelings and thoughts, Forming inner peace by rewards   - Role management: Regaining responsibilities in family and society, Engaging in charitable activity   - Management of social complexities (New Component): Relocating to another environment, Behaving in an assertive manner, Advocacy for social changes. | n/a |
| S5 | Quantitative | Brawley et al. [63]  2013 | Canada | Community |  | Quantitative | Feasibility study | Patients (n=13) | Physical activity | Questionnaire | - Numeric outcome:   - Usability and fidelity measures   - Minutes per week of self-managed and supervised LTPA.   - Self-regulatory efficacy, and action planning - Narrative outcome: n/a | n/a |
| S6 | Qualitative | Cadel et al. [45]  2020 | Canada | Community |  | Qualitative | Qualitative design | Patients (n=19), health professionals (n= 32) | Medication | Interview | - Numeric outcome: n/a - Narrative outcome:   - Themes of strategies: Disease controlling strategies, Process strategies, Resource strategies, Activities strategies, Internal strategies, Social interaction strategies, Self-management support | n/a |
|  | Qualitative | Cadel et al. [46]  2022 | Canada | Community |  | Qualitative | Qualitative design | Patients (n=19) | Medication | Interview | - Numeric outcome: n/a - Narrative outcome:   - Themes of attitudes and beliefs | n/a |
| S7 | Qualitative | Fiordelli et al. [76]  2020 | Switzerland | Community |  | Qualitative | Intervention development | 130 recommendations and 15 experts from the field of SCI and individuals with SCI | Pressure injury | Consensus meeting | - Numeric outcome: n/a - Narrative outcome:   - A set of 98 evidence-based recommendations | n/a |
| S8 | Qualitative | Gourlan et al. [86]  2020 | France | Community |  | Qualitative | Qualitative design | Patients (n=131) | Pressure injury | Open-ended questionnaire | - Numeric outcome: n/a - Narrative outcome:   - Category 1: Identifying what might become problematic.   - Category 2: Daily preventive action.   - Category 3: Detecting the early signs.   - Category 4: Managing the early signs.   - Category 5: Need for care.   - Category 6: Experience with PU and being bedridden | n/a |
| S9 | Quantitative | Guihan et al. [64]  2014 | US | Hospital |  | Quantitative | Experimental design: RCT | Patients (n =6) | Pressure injury | Questionnaire | - Numeric outcome:   - Effectiveness outcome measures: Skin Care Behaviors, Skin Status, Health Care Utilization, Days on Bedrest, Self-Management, PrU Knowledge: - Narrative outcome: n/a | Demographics, SCI  factors, and PrU characteristics.  The Salzburg Risk Score assesses SCI-specific PrU risk factors  Comorbidities were obtained via medical records, Depression through Patient Health Questionnaire, Substance abuse and risky drinking using the Alcohol Use Disorders Identification Test |
| S10 | Qualitative | Guilcher et al. [47]  2020 | Canada | Outpatient setting |  | Qualitative | Qualitative design | Health professionals (n=32) | Medication | Interview | - Numeric outcome: n/a - Narrative outcome:   - Roles of health professionals: Tailoring medications, Providing education, and Exploring alternatives:   - Factors impacting MTM (barriers and enablers): Patient self-management skills, Provider knowledge and confidence, Provider-patient relationships, Interprofessional collaboration, Provider funding models | n/a |
|  | Qualitative | Guilcher et al. [48]  2019 | Canada | Outpatient setting |  | Qualitative | Qualitative design | Health professionals (n=32) | Medication | Interview | - Numeric outcome: n/a - Narrative outcome:   - Micro-level factors: Medication-related factors: Side effects, Perceived effectiveness, Perceived safety, Drug regimen complexity: Complex dosing, timing, and the number of medications could decrease adherence. Providers sometimes simplified regimens to mitigate this issue. Patient-specific factors: Medication knowledge, Patient preferences, expectations, and goals, Severity of injury, cognitive function/mental health, and time since injury, Caregiver support   - Meso-level factors: Provider-specific aspects: Providers' knowledge and confidence, Building patient trust   - Macro-level factors: Health systems factors: Availability and accessibility of healthcare services, Availability and accessibility of medications | n/a |
| S11 | Qualitative | Hirsche et al. [54]  2011 | Canada | Community |  | Qualitative | Qualitative design | Patients (n=22) | Non-specific | Interview | - Numeric outcome: n/a - Narrative outcome:   - Preprogram influences   - Common experiences within the group   - Factors affecting learning opportunities   - Workshop content   - Outcomes | n/a |
| S12 | Quantitative | Hoffman et al. [75]  2023 | US | Community |  | Quantitative | Experimental design: RCT | Patients (n=183) | Non-specific | Questionnaire | - Numeric outcome:   - Quality of life   - Self-efficacy   - Health and participation   - Satisfaction with the program - Narrative outcome: n/a | Age, sex, marital status,  current living situation, socioeconomic status, rural/ urban status, employment and level and completeness  of injury  Depressive symptoms |
| S13 | Quantitative | Houlihan et al. [38]  2017 | US | Community |  | Quantitative | Experimental design: RCT | Patients (n=84) | Non-specific | Questionnaire | - Numeric outcome:   - Health self-management   - Social/role activity limitations   - Global rating of change   - Health-related quality of life   - Communication with physicians   - Patient satisfaction   Narrative outcome: n/a | Age, sex, level of injury, race, education, comorbidity status, years post-injury, income level |
|  | Qualitative | Skeels et al. [39]  2017 | US | Community |  | Qualitative | Experimental design: RCT | Patients (n=84) | Non-specific | Tele-coaching calls interactions | - Numeric outcome: n/a - Narrative outcome:   - Peer health coaches roles | n/a |
| S14 | Qualitative | Jeyathevan et al. [55]  2021 | Canada | In-patient Rehabilitation |  | Qualitative | Qualitative design | Experts in self-management, practitioners, rehabilitation scientists, patient and family educators, partners from community organizations, policy leaders, researchers | Non-specific | Discussions | - Numeric outcome: n/a - Narrative outcome:   - Structure indicator - the proportion of staff with appropriate education and training in self-management principles.   - Process indicator - the proportion of SCI/D inpatients who have received a self-management assessment related to specific patient self-management goal(s) within 30 days of admission.   - Outcome indicator - the Skill and Technique Acquisition, and Self-Monitoring and Insight subscores of the modified Health Education Impact Questionnaire. | n/a |
| S15 | Quantitative | Juengst et al. [65]  2019 | US | Inpatient rehabilitation or acute care and community |  | Quantitative | Feasibility study | Care partners (n=39) | Non-specific | Questionnaire | - Numeric outcome:   - Recruitment Measures: Number and percentage of recruited participants. Reasons for ineligibility. Reasons for refusal.   - Intervention Delivery Measures: Number of PST sessions completed. Length of PST sessions. Pittsburgh Rehabilitation Participation Scale (PRPS) (measuring participant engagement). Intervention uptake. Client Satisfaction Questionnaire-8 (CSQ-8). Working Alliance Inventory (WAI). CaPP   - App Usage Measures: Number of participants who downloaded and used CaPPS. Frequency of on-demand booster session completion. Frequency of weekly booster session completion   Narrative outcome: n/a | n/a |
| S16 | Quantitative | Keegan et al. [66]  2012 | US | Community |  | Quantitative | Experimental study: RCT | Patients (n=126) | Physical activity | Questionnaire | - Numeric outcome:   - Physical activity and exercise participation - Narrative outcome: n/a | Age, gender, race, education, time since injury, preinjury  physical activity level, and functional limitations, employment  status, occupation, income level, preinjury physical activity, Normative and control beliefs, Interpersonal support, Perceived benefits, Perceived barriers, Self-Efficacy, Commitment to a plan |
| S17 | Quantitative | Kooijmans et al. [80]  2017 | Netherlands | Community |  | Quantitative | Experimental design: RCT | Patients (n=64) | Physical activity | Questionnaire | - Numeric outcome:   - Self-reported physical activity   - Amount of self-propelled wheelchair driving   - Perceived behavioral control   - Stages of change concerning exercise   - Attitude toward exercise   - Secondary health complications (SHCs)   - Social support   - Aerobic capacity   - Functional independence   - Mood   - Fatigue   - Participation   - Quality of life   - Body mass index - Narrative outcome: n/a | Age, sex, time since injury, level  of SCI, rehabilitation center, and baseline BMI |
| S18 | Quantitative | Kryger et al. [67]  2019 | US | Community |  | Quantitative | Experimental design: RCT | Patients (n=38) | Urinary infection | Questionnaire | - Numeric outcome:   - Health outcomes: Number of Urinary Tract Infections (UTIs), Number of Pressure Injuries, Number of Emergency Department (ED) Visits, Number of ED Visits Due to UTIs or Pressure Injuries, Number of Hospitalizations, Number of Hospitalizations Due to UTIs or Pressure Injuries   - Psychosocial outcomes: Canadian Occupational Performance Measure (COPM), Adolescent Self-Management and Independence Scale, Beck Depression Inventory-II (BDI-II), Patient Assessment of Chronic Illness Care, World Health Organization Quality of Life Brief Instrument, Craig Handicap Assessment and Reporting Technique Short Form (physical independence domain) - Narrative outcome: n/a | n/a |
| S19 | Quantitative | De Laat et al. [79]  2017 | Netherlands | Community |  | Quantitative | Observational design cross-sectional | Patients (n=170) | Pressure injury | Questionnaire | - Numeric outcome:   - Patient activation measure   - Self-management behaviour statements - Narrative outcome: n/a | Age, gender, marital status, level of education, rehabilitation center, cause of paraplegia, severity, time of paraplegia onset, PU occurrence, PU treatment, physical PU experience, perceived risk of developing a PU,  General Health-Related Factors:  Self-reported health  Quality of life  Global quality of life |
| S20 | Quantitative | Liu et al. [84]  2023 | China | Community |  | Quantitative | Experimental design: RCT | Patients (n=98) | Depression | Questionnaire | - Numeric outcome:   - Depression was assessed using the Beck Depression Inventory-II (BDI-II) at the 12th and 24th weeks post-discharge - Narrative outcome: n/a | Age, sex, education level, marital status, income, occupation, duration of injury, etiology, and American Spinal Injury Association Impairment Scale grade |
| S21 | Quantitative | MacGillivray et al. [56]  2020 | Canada | Rehabilitation hospital and 3 months post discharge |  | Quantitative | Feasibility study | Patients (n=20) | Non-specific | Questionnaire | - Numeric outcome:   - Feasibility measures: Recruitment rate, Retention, Intervention adherence, Physical usability of intervention, Adverse events, Use of and administration of outcome measures - Narrative outcome: n/a | Age, sex, education, employment, living environment, smartphone ownership, and previous experience with mobile health apps, Injury type (Traumatic vs. Non-traumatic),  Neurological level, AIS Grade, Self-Efficacy, Technology Readiness  demographics questionnaire, the Spinal Cord  Independence Measure-III (SCIM-III) self-report version (assesses self-care, respiration, bowel and bladder and mobility), the New General Self Efficacy Scale (NGSES) (measures confidence to deal with daily hassles and stressful life events), the Technology Readiness Index 2.0 (TRI) (captures inclination to adopt new technologies), and a custom Likert scale to identify perceived importance of key  areas of self-management relating to SCI ranging from  1–5 with 1 being very important and 5 being not important. |
| S22 | Quantitative (pilot RCT) | Meade et al. [68]  2016 | US | Community |  | Quantitative (pilot RCT) | Feasibility study | Patients (n=27) | Non-specific | Questionnaire | - Numeric outcome:   - Feasibility outcome measures: recruitment, retention, and completion rates and mode of intervention delivery.   - Effectiveness outcome measures: Spinal Cord Injury Secondary Conditions Scale (SCI-SCS), Patient Health Questionnaire (PHQ-9), Social Problem-Solving Inventory – Revised: Short (SPSI-R:S), Disability Management Self-Efficacy Scale (DMSES), Knowledge measure about managing SCI and preventing secondary conditions - Narrative outcome: n/a | Gender, age, race/  ethnicity, marital status, education, employment  status, and residence, age at injury, time since injury, level and severity of injury, cause of injury, and concurrent injuries |
| S23 | Qualitative | Mortenson et al. [57]  2019 | Canada | In-patient - rehabilitation center |  | Qualitative | Intervention development | Patients (n = 20) and informal caregivers (n = 7) formal caregivers (n = 48) | Non-specific | Interview | - Numeric outcome: n/a - Narrative outcome:   - Themes: Being individualized and user-friendly; Targeting goals to promote self-management; Improving participation and gaining support to facilitate lifestyle change | n/a |
| S24 | Qualitative | Munce et al. [49]  2014 | Canada | Community |  | Qualitative | Qualitative design | Patients (n=7), caregivers (n=7), rehabilitation managers (n=12) | Non-specific | Interview | - Numeric outcome: n/a - Narrative outcome:   - Facilitators to self-management: Physical support from the caregiver, Emotional support from the caregiver, Peer support and feedback, Importance of positive outlook and acceptance, Maintaining independence/control over care   - Barriers to self-management: Caregiver burnout, Funding and funding policies, Lack of accessibility, Physical limitations and secondary complications, Difficulties achieving positive outlook or mood | n/a |
|  | Qualitative | Munce et al. [50]  2016 | Canada | Community |  | Qualitative | Qualitative design | Patients (n=7), family/caregivers (n=7), acute care/rehabilitation managers (n=12) | n/a | Interview | - Numeric outcome: n/a - Narrative outcome:   - Internal responsibility attribution: Wellness awareness, Monitoring for secondary complications, Independence-dependence conflict, Directing someone else to provide your care, Ownership of your own care/empowerment   - External responsibility attribution: Established chronic disease self-management programs, Importance of caregiver skill set | n/a |
| S25 | Quantitative | Munce et al. [58]  2014 | Canada | Community |  | Quantitative | Observational design cross-sectional | Patients (n=99) | Non-specific | Questionnaire | - Numeric outcome:   - Importance ratings of self-management program components.   - Preferences regarding the delivery format, construction of program components, timing, follow-up periods, program leaders, and program organizers - Narrative outcome: n/a | Age, gender, marital status, level of education, level of injury, and time since injury |
| S26 | Qualitative | Munce et al. [59]  2017 | Canada | Community |  | Qualitative | Qualitative design | Patients (n=3), clinicians (n=5), researchers (n=14), policy makers (n=3) | n/a | Brainstorming  Sessions | - Numeric outcome: n/a - Narrative outcome:   - Themes: Knowledge domain, Skills domain, Social/professional role and identity domain, Beliefs about capabilities domain, Beliefs about consequences domain, Reinforcement, Intentions, and goals domains, Memory, attention, and decision processes domain, Environmental context and resources domain, Social influences domain, Optimism/emotion domain, Behavioral regulation domain | n/a |
| S27 | Quantitative | Newman et al. [69]  2019 | US | Community |  | Quantitative | Feasibility study | Patients (n=10) | Pressure injury, urinary infections, bowel management | Observation and questionnaire | - Numeric outcome:   - Usability and acceptability of iPad   - Usability and acceptability of iTunes U and course content   - Usability and acceptability of video chat (FaceTime) - Narrative outcome: n/a | n/a |
| S28 | Qualitative | Oh et al. [70]  2023 | US | Community |  | Qualitative | Qualitative design | Patients (n=5) | Skin management | Interviews and questionnaire | - Numeric outcome:   - Participants' adherence to pressure relief (PR) exercises, measured through self-reported completion of PRs in response to reminders sent by the chat-based mobile probe - Narrative outcome:   - Unique experiences to similar problems   - Increasing PR adherence through a personalized experience   - Gamifying and visualizing PR adherence | n/a |
| S29 | Mixed-methods | Olney et al. [71]  2019 | US | Community |  | Mixed-methods | Feasibility study | Patients (n=18) | Pressure injury | Survey and interview | - Numeric outcome:   - App usability - Narrative outcome:   - Themes, suggestions, and feedback   - Integration of components   - The value of real-time pressure mapping   - Privacy concerns | n/a |
| S30 | Qualitative | Pancer et al. [60]  2019 | Canada | Community |  | Qualitative | Qualitative design | Patients (n=13), healthcare professionals (n=9) | Physical activity | Interview | - Numeric outcome: n/a - Narrative outcome:   - Preferred features: Behavior change techniques   - Knowledge: Guidance, Barrier management,   - Possibility of achievement: Risks and benefits, Modelling   - Self-Regulation strategies: Action Planning, Goal Setting, Tracking, Rewards , Reminders   - Modes of delivery   - Interactivity: Peer, Professional   - Format: Appearance, Language, Ease of Use | n/a |
| S31 | Qualitative | Pilusa et al. [87]  2021 | South Africa | Community |  | Qualitative | Qualitative design | Patients (n=17) | Non-specific | Interview | - Numeric outcome: n/a - Narrative outcome:   - Themes: Prevention of secondary health conditions; Management of secondary health conditions; and Challenging experiences | n/a |
| S32 | Quantitative | Potiart et al. [83]  2020 | Thailand | Inpatient rehabilitation |  | Quantitative | Observational design cross-sectional | Patients (n=35) | Urinary catheter | Survey | - Numeric outcome:   - User satisfaction   - Usability   - Catheter-Related Pain   - Leakage   - Urinary Tract Infection - Narrative outcome: n/a | n/a |
| S33 | Qualitative | Pryor et al. [88]  2021 | Australia | Community |  | Qualitative | Qualitative design | Patients (n=11) | Bowel care | Interview | - Numeric outcome: n/a - Narrative outcome:   - Usual bowel care practices: Bowel care regimens, Assistance, Frequency and timing, Integrating bowel care into everyday life: Participants emphasized the critical role of effective bowel care in their daily lives, impacting physical health, well-being, and the prevention of accidents and complications.   - Four key factors for integration of bowel care: Acceptance, motivation, and willingness, Discipline, Proactive self-management, Fostering collaboration with carers | n/a |
| S34 | Quantitative | Raghavan et al. [90]  2003 | UK | Community |  | Quantitative | Observational design cross-sectional | Patients  (n=472) | Pressure injury | Questionnaire | - Numeric outcome:   - Point prevalence of pressure sores - Narrative outcome: n/a | Inspection of skin for pressure damage at least once a day.  Lifting body weight while seated at least once an hour.  Fecal incontinence at least few times a week  Urinary incontinence at least few times a week.  Smoking at the time of survey.  Concurrent medical problems.  Gender  Neurological level  Living alone  Employed  Age |
| S35 | Qualitative | Shirai et al. [61]  2022 | Canada | Community |  | Qualitative | Qualitative design | Patients (n=9) | Pressure injury | Interview | - Numeric outcome: n/a - Narrative outcome:   - Strengths and weaknesses of PUT: Benefits and usefulness, Potential challenges and pitfalls, Navigation, Aesthetics   - Target population for PUT: Varied characteristics of users, Timing post-injury   - Key concepts and messages as motivators for using PUT: Prevention, Impact on quality of life   - Recommendations for improvement of PUT: Inclusion of additional information, Changes to visuals and aesthetics, Changes to accessibility | n/a |
| S36 | Mixed-methods | Singh et al. [62]  2020 | Canada | Inpatient rehabilitation |  | Mixed-methods | Feasibility study | Patients (n=20) | Non-specific | Survey and user feedback | - Numeric outcome:   - Usability - Narrative outcome   - Themes: Being accessible to users, Being intuitive to navigate, Offering users flexibility. | n/a |
| S37 | Qualitative | Starosta et al. [74]  2024 | US | Community |  | Qualitative | Qualitative design | Patients (n=158) | Non-specific | Online forum posts | - Numeric outcome: n/a - Narrative outcome:   - Themes: Skill building, Resource sharing, Problem solving, Bearing witness | n/a |
| S38 | Qualitative | Van Gaal et al. [81]  2022 | Netherlands | Community |  | Qualitative | Qualitative design | Patients (n=14) | Pressure injury | Interview | - Numeric outcome: n/a - Narrative outcome:   - Managing the medical tasks of prevention or treatment of pressure ulcers: Managing preventing pressure ulcers, Managing various types of support, Managing treatment of pressure ulcers   - Managing high-risk situations   - Managing the emotional impact of preventing and treating pressure ulcers: Dependence on others, Hospitalization, Honesty and support   - Managing the prevention and treatment of pressure ulcers in social, everyday life: Managing family life, Managing work and hobbies | n/a |
| S39 | Quantitative | Wang et al. [85]  2023 | China | Community |  | Quantitative | Observational design cross-sectional | Patients (n=110) | Skin management | Questionnaire | - Numeric outcome:   - Revised skin management needs assessment checklist (Revised SMnac) - Narrative outcome: n/a | Age, gender, education level, medical expense reimbursement ratio, etiology, level of lesion, injury severity, time since injury, comorbidity, knowledge about skin self-management, attitude, self-efficacy, Spinal Cord Independence Measure |
| S40 | Qualitative | Widerström et al. [73]  2023 | US | Community |  | Qualitative | Qualitative design | Patient (n=15), significant other (n=12), health professionals (n= 10) | Neuropathic pain | Interview | - Numeric outcome: n/a - Narrative outcome:   - Content   - Comprehensibility   - Format | n/a |
| S41 | Qualitative | Wilde et al. [40]  2011 | US | Community |  | Qualitative | Qualitative design | Patients (n=34) | Urinary catheter | Questionnaire and interviews | - Numeric outcome: n/a - Narrative outcome:   - Themes: Knowing the body, Practicing clean intermittent catheterization (CIC), Limited options in catheters and equipment, Inaccessible bathrooms, Hassles, Adjustment in making CIC a part of life   - Catheter practices, frequency of urinary tract infection, leakage and pain | n/a |
|  | Qualitative | Wilde et al. [41]  2015 | US | Community |  | Qualitative | Intervention development | Patients (n=34) | Urinary catheter | Discussion | - Numeric outcome: n/a - Narrative outcome:   - Components: Educational Materials, Personal Data Section   - Study Nurse Consultations: Peer-Led Online Forum Discussions, Mobile Phone Modification | n/a |
|  | Mixed-methods | Wilde et al. [42]  2016 | US | Community |  | Mixed-methods | Feasibility study | Patients (n=34) | Urinary catheter | Questionnaire and interview | - Numeric outcome:   - Self-management scores   - Feasibility   - Usability   - Urinary diary usage   - Self-efficacy and HRQOL - Narrative outcome:   - Changes in self-care management | n/a |
| S42 | Qualitative | Zanini et al. [77]  2020 | Switzerland | Community |  | Qualitative | Qualitative design | Patients (n=20) | Pressure injury | Interview | - Numeric outcome: n/a - Narrative outcome:   - Styles of prevention: Thoughtfuls, Selectives, Delegators   - Knowledge and attitude regarding prevention: Extensive vs. Basic Knowledge, Personal responsibility vs. Delegation, Priority of prevention   - Attitudes toward life with SCI: Optimism, Self-Efficacy, Proactivity | n/a |
| S43 | Qualitative | Zanini et al. [78]  2019 | Switzerland | Inpatient and community |  | Qualitative | Qualitative design | Health professionals (n=26) | Pressure injury | Interview | - Numeric outcome: n/a - Narrative outcome:   - Themes: Defining responsibilities and expectations, Negotiating priorities and setting common goals, Building a basis of mutual trust and respect | n/a |
| S44 | Mixed-methods | Zhou et al. [72]  2020 | US | Community |  | Mixed-methods | Feasibility study | Patients (n=5) | Non-specific | Questionnaire  and interview | - Numeric outcome:   - Accessibility improvement   - Usability - Narrative outcome   - Performance on tasks | n/a |
