## Supplemental Table 3 for "Trends in self-management research in spinal cord injury: A scoping review of study designs and findings"

| Author | Objective | Synthesized results | Self-management tasks as per Lorig et al.'s [32] |
| --- | --- | --- | --- |
| Allin et al.  [43]  2020 | To evaluate the feasibility and potential impact of the spinal cord injury program in the context of a mixed methods pilot study. | The program exhibited high feasibility, with an 81% retention rate (10 out of 11 participants completed the program) and 100% adherence, as all participants completed all sessions. Sessions had a median duration of 55 minutes, with most participants completing them within 14 days of each other. Notably, participants frequently opted to use familiar communication tools (e.g., Zoom, Skype) rather than the built-in videoconferencing tool. The program covered a range of health management topics, including diet, exercise, mental health, bowel and bladder care, skin health, and sexuality.  Positive changes were observed across all measured outcomes, including self-efficacy, resilience, physical health status, emotional status, and health literacy. While the changes were not statistically significant, they were consistently positive. The most notable improvements were in self-efficacy and electronic health literacy, both of which showed moderate effect sizes. Smaller effect sizes were observed for resilience, depression, and secondary conditions, indicating modest but meaningful progress in these areas.  Interviews revealed that participants experienced various benefits from the program, including improvements in diet, exercise, and mental health. Health coaches played a critical role in the program's success, serving as sources of accountability, inspiration, health information, social support, and technical assistance. Participants highly valued the coaches' supportive roles, describing them as motivators, role models, and troubleshooters who enhanced their overall experience. | Medical management  Emotional management |
| Allin et al.  [44]  2018 | To provide details of a participatory design process for an internet-mediated self-management program for users with SCI and illustrate how it has been used to define design constraints and solutions. | Participants emphasized the critical role of the internet in self-management, noting its utility in discovering services and interventions, interacting with supportive communities, locating specific community services, preparing for healthcare appointments, revisiting rehabilitation skills, accessing research information, and reducing the burden of travel. However, a review of existing online resources revealed several challenges: 1) Lack of familiarity: Participants were often unfamiliar with online resources, such as health information forums. 2) Appreciation of diversity: Participants valued the diversity of self-management resources available online, recognizing that different resources might suit different individuals. 3) Questions of trust: Concerns about the credibility of online information were prevalent, with participants preferring advice from trusted sources like healthcare professionals and peers. 4) Accessibility concerns: Participants highlighted accessibility issues, noting that many online resources were not tailored to the specific needs of individuals with SCI.  Participants proposed innovative solutions to address the challenges identified during the exploration phase: 1) Community-curated resource database: Participants suggested creating a collaborative, Wiki-like database to filter and lend credibility to online self-management resources. 2) Online information navigators: The idea of internet-accessible information navigators was proposed to guide users and filter useful information from online collectives. 3) Online groups: Participants found value in regular peer and researcher meetings during the exploration phase, which enhanced their awareness of self-management strategies and techniques.  In the prototyping phase, participants discussed mechanisms to ensure the credibility and usability of the proposed database: 1) Stars and up-voting: Participants recommended using community ratings (e.g., stars) and up-voting/down-voting systems to identify credible and valuable information. 2) Expertise points and activity feeds: While the idea of rewarding contributors with "points" based on the value of their contributions was considered, it was seen as controversial. Instead, participants favored activity feeds on user profiles to showcase contributions without fostering competition. | Medical management  Emotional management |
| Amann et al. [36]  2020 | To (1) establish a co-design approach for developing a high-fidelity prototype app for the self-management of individuals with spinal cord injury, (2) design the prototype that resulted from this process, and (3) conduct the first usability assessment of the prototype app. | The prototype was tested by 15 participants through task completion exercises. The results demonstrated a high level of usability, with most participants able to complete tasks with ease, indicating that the app was intuitive and functional for its intended purpose. Participants generally praised the app's logical and intuitive structure, describing it as simple, user-friendly, and easy to navigate. However, they provided constructive feedback to enhance the app further, including: The need for larger font sizes to improve readability; An overview of all app features to help users understand the full range of functionalities; A search function to allow users to locate relevant content more efficiently. Some participants experienced minor navigation difficulties, which were attributed to the testing device (Android) rather than the app's design. Participants more familiar with iPhones found it slightly challenging to adapt to the Android interface during testing. Several participants noted that initial navigation challenges were part of the natural learning process when using a new app. They expressed confidence that users would become more comfortable with the app over time through trial and error. | Medical management |
| Amann et al. [37]  2020 | To identify the perceived benefits of a co-designed self-management app that could promote its uptake and to explore the factors that may impede adoption. | Benefits that promoted the app uptake were: A Companion for newly injured individuals: Both wheelchair users and healthcare professionals recognized the app's value for individuals who had recently experienced a spinal cord injury, particularly during initial rehabilitation. The app was seen as a complementary tool to traditional patient education materials, offering interactive and customizable information that could engage users more effectively. It empowered patients to learn at their own pace, fostering greater autonomy. An emergency kit and motivational support: Participants highlighted the app's utility in extraordinary situations, such as during travel or outside regular healthcare service hours. It was viewed as a valuable resource for contacting healthcare professionals or obtaining advice when immediate assistance was unavailable. The app's smart camera feature was particularly praised for its potential to assess early-stage pressure injuries, providing timely and practical support. A guide for informal caregivers and family members: Informal caregivers and family members were identified as key beneficiaries of the app. It could provide them with curated, evidence-based information about SCI and pressure injury prevention, helping them better understand and support individuals with SCI. This feature was seen as a way to extend the app's impact beyond the primary user.  The challenges impeding app adoption were: Motivating individuals to use the app: A significant challenge was motivating individuals, both experienced and newly injured, to use the app proactively for preventive purposes. Some wheelchair users felt they had already internalized preventive behaviors and might not need reminders. There was concern that users might only turn to the app reactively, after experiencing skin problems, rather than using it as a preventive tool. Concerns about misuse and abuse of the app: Participants raised concerns about potential misuse, such as individuals contacting healthcare professionals for irrelevant information or social interaction. This could lead to an overload of messages, potentially overwhelming healthcare providers and reducing the app's effectiveness. Organizational and maintenance challenges: Integrating the app into existing healthcare systems and workflows posed logistical challenges. Determining eligibility criteria for app use and managing resources were organizational concerns. Additionally, maintaining the app's content with up-to-date, evidence-based information was seen as critical but raised questions about responsibility and resource allocation. | Medical management |
| Baniya et al. [82]  2022 | To examine the prevalence, severity, and self-management of depressive mood in community-dwelling people with spinal cord injury. | Of the 115 participants, 84.3% reported experiencing a depressive mood, and 60.8% of these individuals exhibited moderate to severe levels of depression. To manage their depressive symptoms, participants primarily turned to non-pharmacological strategies, such as using the internet and social media, sharing feelings with family members, engaging in Hindu religious practices, substance abuse (e.g., alcohol, cannabis), and crying. These methods were chosen for their ease of use, ability to provide a sense of relaxation and peacefulness, and the lack of accessible alternative comfort measures. Among these strategies, sharing feelings, using the internet and social media, and participating in Hindu religious practices were reported as the most effective in alleviating depressive symptoms.  While many of the strategies were perceived as helpful, the reliance on substance abuse as a coping mechanism points to the urgent need for accessible mental health support and alternative interventions tailored to this population. | Medical management  Emotional management |
| Begum et al. [89]  2022 | To explore how community-dwelling persons with spinal cord injury and their primary caregivers execute self-management strategies in daily activities. | The research identified nine groups of self-management strategies, some of which extended beyond the traditional components of medical, emotional, and role management. As a result, a new component, termed "management of social complexities," was proposed to better capture the broader challenges faced by individuals with SCI. Medical management: Participants emphasized the importance of developing and maintaining consistent routines and habits, such as changing positions to prevent pressure ulcers and adhering to a healthy diet, as advised by healthcare professionals. Additionally, they highlighted the health-promoting properties of daily activities, such as engaging in physical movements, which helped prevent complications like pressure ulcers and joint pain. Emotional management: To cope with the emotional consequences of SCI, participants discussed strategies such as developing authority over their feelings and thoughts through recreational activities, positive thinking, and maintaining a sense of identity and purpose. Achieving personal or societal rewards, such as respect and recognition for their contributions, also played a significant role in fostering inner peace and emotional well-being. Role management: Participants focused on regaining responsibilities within their families and communities, either by resuming previous roles or creating new ones. Engaging in charitable activities was another key strategy, allowing individuals to become role models and make positive contributions to their communities, thereby enhancing their sense of purpose and belonging.  Management of social complexities (New component): This newly proposed component addressed the broader social challenges faced by individuals with SCI. Strategies included relocating to more disability-friendly environments to escape negative attitudes and discrimination, which enabled greater participation in daily activities. Participants also demonstrated assertiveness in advocating for their interests, such as property division, and worked to change societal perceptions of individuals with SCI. Additionally, they engaged in advocacy efforts to promote accessibility and defend their rights, often seeking support from organizations to address barriers to participation. | Medical management  Emotional management  Role management |
| Brawley et al. [63]  2013 | To test the efficacy and feasibility of a group-mediated cognitive-behavioral (GMCB) intervention to increase self-managed leisure time physical activity (LTPA) | Increase in LTPA: Participants demonstrated a significant increase in LTPA, nearly doubling their total minutes per week from an average of 42.00 ± 69.57 minutes at baseline to 197.50 ± 270.86 minutes post-intervention (p < .05). This increase was attributed to the self-managed LTPA plans developed during the intervention, which did not interfere with the time spent in supervised LTPA. Self-regulatory efficacy: The intervention successfully sustained participants' self-regulatory efficacy, with scores remaining high from baseline (M = 86.20 ± 10.49) to post-intervention (M = 89.43 ± 10.23). This indicates that participants maintained their confidence in managing their LTPA independently. Action planning: Action planning showed a near-significant improvement, increasing from baseline (M = 4.63 ± 3.25) to post-intervention (M = 6.83 ± 2.40; p = .06). This suggests that participants became more adept at creating and implementing plans for their LTPA. Usability and fidelity: The intervention materials and protocol were perceived as highly usable by both participants and the interventionist, indicating strong intervention fidelity. This reflects the practicality and effectiveness of the program's design and delivery. Satisfaction and learning outcomes: Participants reported a significant increase in their perceived likelihood of obtaining physical benefits from added LTPA (p = .04). They also provided positive feedback on the learning outcomes and intervention content, highlighting the program's success in enhancing their understanding and motivation for LTPA. | Medical management |
| Cadel et al. [45]  2020 | To explore attitudes and experiences of medication self-management from the perspectives of persons with spinal cord injury/dysfunction and providers, and to explore the extent to which the Taxonomy of Everyday Self-management Strategies framework captured participants' experiences with medication self-management. | Participants employed various strategies to manage their medications, including disease-controlling strategies such as balancing symptom management with side effects, though some individuals stopped medications due to unwanted side effects. Process strategies involved problem-solving and decision-making to address side effects, adjust medication routines, and actively participate in treatment decisions, with healthcare providers stressing the importance of self-advocacy. Resource strategies included seeking support from healthcare providers, advocating for drug coverage, and relying on information from professionals or unpaid help from family and friends. Activities strategies focused on organizing medication-taking using tools like planners, alarms, and apps, while adjusting schedules to fit meaningful daily activities. Internal strategies, though less discussed, revealed challenges in accepting medication use post-injury, with some viewing it as a sign of weakness. Social interaction strategies were rarely mentioned, though community involvement and maintaining connections with loved ones were highlighted as important for emotional well-being.  Healthcare providers played a vital role in supporting medication self-management by teaching self-advocacy, educating patients about medications, and fostering shared decision-making. | Medical management |
| Cadel et al. [46]  2022 | To explore the attitudes, beliefs and experiences pertaining to the management of prescribed and unprescribed medications among community-dwelling adults with spinal cord injury/dysfunction. | Participants, many of whom had not taken medications prior to their injury, found the sudden need for complex medication regimens disruptive. Managing multiple medications with specific timing and limited knowledge about the drugs posed significant challenges. Participants described medications as an "essential evil," expressing tension between their desire for independence and the necessity of taking medications to manage their health. Over time, they came to appreciate the benefits of medications, such as improved function and relief from secondary complications, despite initial reluctance.  The effectiveness of medications varied among participants, with some finding their regimens effective in managing secondary complications, while others did not achieve the desired relief. Many participants reported experiencing side effects, such as fatigue, drowsiness, dry mouth, and impaired memory, which disrupted their daily lives. Safety concerns related to long-term medication use and potential adverse effects on various body organs were also raised, though participants often felt compelled to continue taking medications due to their perceived necessity. Fear of negative outcomes, such as debilitating secondary complications or side effects, made participants hesitant to adjust or add medications.  Self-management played a crucial role in medication management. Participants developed individualized strategies, such as establishing daily routines or using medication packaging (dosettes), to ensure they took their medications correctly. Some participants initiated medication adjustments without clinical supervision, driven by personal preferences and experiences, while others made adjustments under clinical guidance. | Medical management |
| Fiordelli et al. [76]  2020 | To present a procedure for the participatory identification of evidence-based content to ground the development of a self-management app. | The process involved three key stages. In Stage 1, an environmental scan was conducted to review and categorize existing recommendations related to PI prevention and self-management in SCI. A total of 130 recommendations were identified from various sources and organized into 12 topics, such as support surfaces, repositioning, nutrition, and skin care. Stage 2 involved a two-part consensus meeting with SCI experts and individuals with SCI. In Part I, 15 recommendations were excluded, 60 were included, and 55 were deemed ambiguous. Part II addressed the ambiguous recommendations, resulting in the exclusion of nine and the inclusion of 25. Additionally, 62 recommendations required further specification, and experts provided additional details and criteria for these. In Stage 3, the research team reviewed the remaining ambiguous recommendations, including seven and excluding 14. A working group of nutritionists and SCI-specialized medical doctors developed six new nutrition recommendations. The final outcome was a comprehensive set of 98 evidence-based recommendations for the prevention and management of PIs in community-dwelling individuals with SCI. | Medical management |
| Gourlan et al. [86]  2020 | To explore the perceptions and beliefs related to pressure ulcers, their prevention and treatment strategies, in order to discuss potential learning objectives for pressure ulcers-related therapeutic education in persons with spinal cord injury. | Participants recognized factors contributing to PU formation, such as prolonged seating, muscle atrophy, and skin aging, and acknowledged their own susceptibility due to physical, psychological, or environmental factors. Daily preventive actions included reliance on caregivers for monitoring, the use of specialized equipment (though sometimes blamed for PU), and varying levels of attention to skin care and physical measures. Integrating prevention into daily life was often seen as time-consuming and restrictive. Detecting early signs of PU involved skills like identifying pain or vegetative signs, though challenges such as skin anesthesia and difficulty visualizing at-risk areas were common. Managing early signs included actions like pressure relief and increased vigilance, though some lacked effective strategies. The need for care often led to difficulties in trusting new healthcare providers and evoked negative feelings about returning to hospitals for treatment. Finally, the experience of PU and bed rest had significant psychological and social impacts, including anxiety, depression, social withdrawal, and isolation, with participants emphasizing these consequences over health risks. | Medical management |
| Guihan et al. [64]  2014 | To compare a multicomponent motivational interviewing self-management intervention with a multicomponent education intervention to improve skin-protective behaviors and prevent skin worsening in veterans with spinal cord injury hospitalized for severe pressure ulcers. | Effectiveness was measured through skin care behaviors, skin status (documented with digital photographs and planimetry), health care utilization, days in bedrest, self-management, and knowledge.  More than half of the participants experienced skin worsening, with no significant differences between the SM+MI and education control arms at 3 or 6 months. Similarly, skin-related visits and admissions did not differ between the two groups. Baseline demographic, medical, SCI, and ulcer characteristics were comparable across both arms, and most participants in both groups received a minimally effective dose of at least 4 calls, with no significant difference in adherence. Fidelity to MI techniques and spirit was significantly better in the SM+MI arm compared to the education control arm. While the primary outcome, self-reported improvement in skin care behaviors, showed greater improvement in the SM+MI arm at 3 and 6 months, these differences were not statistically significant. Other secondary outcomes, including health care utilization and days in bedrest, also did not show significant differences in intention-to-treat analyses.  Overall, the study found no significant differences in skin worsening, health care utilization, or most secondary outcomes between the SM+MI and education control interventions. However, the SM+MI arm demonstrated better fidelity to MI techniques, suggesting potential for further refinement and investigation of this approach in future studies. | Medical management |
| Guilcher et al. [47]  2020 | To explore healthcare and service providers experiences with medication therapy management for persons with spinal cord injury/disease. | The findings highlighted the distinct roles of different professions in facilitating MTM, with shared tasks such as tailoring medications, providing education, and exploring alternatives. While most providers perceived their care for SCI/D patients as similar to that for other populations, they acknowledged unique challenges related to the physical limitations and medical complexity of SCI/D. Several factors influenced MTM effectiveness. Patient self-management skills, including knowledge of SCI/D-specific medications and information-seeking behaviors, were identified as both barriers and enablers. Providers' lack of knowledge about SCI/D and its medications affected their confidence in supporting MTM, while positive provider-patient relationships and trust facilitated better outcomes. Interprofessional collaboration, whether within multidisciplinary teams or through external consultations, was critical for effective MTM. However, current funding models for pharmacists and physicians were seen as barriers, limiting the time and resources available for comprehensive MTM. | Medical management  Role management |
| Guilcher et al. [48]  2019 | To explore healthcare providers' conceptualization of factors impacting medication adherence for persons with spinal cord injury/disease. | At the micro level, medication-related factors such as intolerable side effects, perceived effectiveness, safety concerns, and drug regimen complexity influenced adherence. Side effects often led patients to alter regimens without consulting providers, while perceived symptom improvement and simplified regimens promoted adherence. Patient-specific factors, including medication knowledge, preferences, injury severity, cognitive function, mental health, time since injury, and caregiver support, also played significant roles. Well-informed patients and strong caregiver support enhanced adherence, whereas cognitive impairments, mental health issues, and patient preferences for avoiding medications posed challenges. At the meso level, providers' knowledge and confidence in managing SCI/D and related medications indirectly impacted adherence by shaping the quality of patient education and trust-building. Establishing trust was crucial for improving information exchange and motivating patients to adhere to their regimens. At the macro level, health system factors such as the availability and accessibility of healthcare services and medications significantly influenced adherence. Challenges included transportation difficulties, long wait times, inaccessible facilities, and limited government funding for services. Providers adapted by offering virtual appointments. Medication cost, insurance coverage, refill policies, and home delivery services also affected adherence, with prohibitive costs and inadequate coverage reducing adherence, while supportive policies and delivery services facilitated it. | Medical management  Role management |
| Hirsche et al. [54]  2011 | To explore the experience of people with neurological conditions who take the chronic disease self-management programme. | Participants' experiences were influenced by pre-program factors such as notifications about the program, motives for enrollment, and expectations. Many joined seeking a supportive environment with peers facing similar challenges and opportunities to learn new information. The group setting was generally positive, fostering understanding and connection, though opinions varied on whether groups should include individuals with similar or different conditions. Younger participants often desired more age-specific peers. The timing of the program's delivery was perceived differently; stroke survivors preferred earlier participation, while those with spinal cord injury (SCI) needed more time to adjust. The program's delivery, including facilitators and workshop format, was appreciated, though some felt it was not adequately adapted for SCI participants. Workshop content, particularly goal setting, was highlighted as a key component, helping participants break down goals into manageable steps. Coping strategies and self-management tools were valued, though some found the information too basic, and the need for condition-specific content varied. Outcomes of the program included positive behavior changes such as smoking cessation, improved eating habits, and increased exercise. Participants gained a stronger sense of self-management and empowerment, valuing the social connections and confidence built through helping others in the group. | Medical management |
| Hoffman et al. [75]  2023 | To assess the impact of a peer-led online self-management program (SCI Thrive) on individuals with spinal cord injury. | While no significant differences were found between the treatment and waitlist groups for primary outcomes such as quality of life, self-efficacy, and life space assessment, several notable improvements were observed within the treatment group. Occupational functioning significantly improved in the treatment group compared to the waitlist group (6.25 vs. -4.07, P = .02). Self-efficacy showed significant improvement from baseline to 6 weeks (6.32 to 6.81, P < .001) and was maintained at 3 months (6.83, P = .001) among those who completed the program. Higher engagement in SCI Thrive was associated with better quality of life (P = .001), increased self-efficacy (P = .007), and greater mobility (P = .026). The COVID-19 pandemic impacted outcomes, particularly mobility and quality of life, due to restrictions and changes in participants' activities. However, participants valued the program for providing stress management tools and fostering connections during the pandemic. Satisfaction with SCI Thrive was high, with 72.6% of participants rating it as "Excellent" or "Very Good." Additionally, 95.9% found the course relevant and useful, and 97.3% would recommend it to others with SCI. | Medical management |
| Houlihan et al. [38]  2017 | To evaluate the impact of My Care My Call" (MCMC), a peer-led, telephone-based health self-management intervention in adults with chronic spinal cord injury. | The primary outcome, health self-management, measured by the Patient Activation Measure (PAM), showed significant improvements in intervention participants at 4 and 6 months compared to the control group. Exploratory subgroup analyses revealed that the intervention was particularly effective for individuals with high social support, those 1 to 6 years post-injury, individuals with tetraplegia, males, white participants, and those with higher education levels. Secondary outcomes indicated that intervention participants experienced significantly greater reductions in social/role activity limitations, increased life satisfaction, improved awareness of services and resources, and greater overall service use at 6 months compared to controls. However, there were no significant changes in communication with physicians or patient satisfaction between the intervention and control groups. | Medical management |
| Jeyathevan et al. [55]  2021 | To describe the selection of self-management structure, process and outcome indicators for adults with spinal cord injury/disease in the first 18 months after rehabilitation admission. | The structure indicator focused on the education and training of rehabilitation staff in self-management principles. It was assessed using a self-assessment tool completed by healthcare professionals (HCPs) or peer mentors, which evaluated their training, skills, comfort, and confidence in providing self-management support to individuals with SCI/D.  The process indicator measured the proportion of SCI/D inpatients who received a self-management assessment related to specific patient goals within 30 days of admission. The Self-Management Working Group refined the SCI-SMET (Spinal Cord Injury Self-Management Evaluation Tool) to assess constructs such as goal setting, problem-solving, action planning, and self-monitoring. The SCI-SMET was pilot-tested in an outpatient clinic, where patients completed the tool in an average of 7-10 minutes. Challenges during pilot testing included difficulties related to cognitive and motor deficits in self-administering the tool and the need for clearer language in the tool.  The outcome indicator assessed the acquisition of self-management skills by individuals with SCI/D using the Health Education Impact Questionnaire (heiQ). The heiQ, a psychometrically validated tool, includes eight dimensions, with "Skill and Technique Acquisition" and "Self-Monitoring and Insight" identified as particularly relevant for SCI/D. Patients rated their confidence in completing specific tasks on a 5-point Likert scale, providing insights into their self-management abilities. | Medical management |
| Juengst et al. [65]  2019 | To determine the feasibility of delivering an evidence-based self-management intervention, problem-solving training, to care partners of individuals with traumatic ain injury, spinal cord injury, burn injury, or stroke during the inpatient imhere  . | Care partners received up to six PST sessions, either in person or via telephone, focusing on a metacognitive strategy to address self-selected problems. Recruitment measures indicated the number and percentage of participants recruited, reasons for ineligibility, and reasons for refusal. Intervention delivery measures included the number and length of PST sessions completed, participant engagement rated using the Pittsburgh Rehabilitation Participation Scale (PRPS), and intervention uptake assessed through participant confidence in applying the PST strategy. Participant satisfaction was measured using the Client Satisfaction Questionnaire-8 (CSQ-8) at one month post-discharge, and the perceived working alliance with the interventionist was evaluated using the Working Alliance Inventory (WAI). Additionally, the study tracked the usage of the CaPPS app, including the number of participants who downloaded and used it, as well as the frequency of on-demand and weekly booster sessions. Participants reported high satisfaction with the intervention, with a mean satisfaction score of 3.35 (on a 1-4 scale). Interventionists rated participants who completed at least one PST session as having very good engagement, with a mean score of 4.75 (on a 1-6 scale). Participants expressed confidence in applying the PST strategy after an average of 2.6 sessions and reported a strong working alliance with interventionists, with a mean WAI score of 6.8 (on a 0-7 scale). Some participants engaged with the CaPPS app's booster sessions multiple times, suggesting the feasibility of using a smartphone app to complement the behavioral intervention for care partners. | Medical management |
| Keegan et al. [66]  2012 | To validate Pender's Health Promotion Model (HPM) as a motivational model for exercise/physical activity self-management for people with spinal cord injuries. | The final model explained 41% of the variance in physical activity and exercise participation, representing a large effect size. Key predictors of participation included preinjury physical activity levels, severity of SCI, and commitment to a plan of action, with commitment emerging as the strongest predictor.  A secondary analysis focused on predicting commitment to a plan of action, a critical behavioral outcome for physical activity participation. Friend and family support, perceived benefits of exercise, and perceived self-efficacy were significant predictors of commitment, while control beliefs did not contribute significantly to the model. | Medical management |
| Kooijmans et al. [80]  2017 | To evaluate the effectiveness of a structured self-management intervention to promote an active lifestyle in inactive persons with long-term spinal cord injury. | Primary outcomes, including self-reported physical activity and the amount of self-propelled wheelchair driving, showed no significant differences between the intervention and control groups. Similarly, no significant changes were observed in perceived behavioral control, which included exercise self-efficacy and proactive coping. However, a positive trend was noted in the stages of exercise change, favoring the intervention group, while exercise attitude yielded mixed results between the groups. At the tertiary level, the intervention group experienced significantly fewer secondary health complications (SHCs) at one of the follow-up assessments compared to the control group. Other tertiary outcomes, such as aerobic capacity, functional independence, mood, fatigue, participation, quality of life, and body mass index (BMI), did not show significant differences between the groups.  Overall, while the intervention did not significantly impact primary or most secondary outcomes, the reduction in secondary health complications and the positive trend in stages of exercise change suggest potential benefits of the HABITS program. | Medical management |
| Kryger et al. [67]  2019 | To determine if the use of iMHere would be associated with improved health outcomes over a 9-month period. A secondary objective was to determine if the use of iMHere would be associated with improved psychosocial outcomes. | Participants in the intervention group experienced a significant reduction in the number of UTIs during the study period compared to before the intervention. However, no significant changes were observed in other primary health outcomes, such as pressure injuries, ED visits, or hospitalizations, in either the intervention or control groups. Additionally, there were no statistically significant differences in psychosocial outcomes, including quality of life, depression, or self-management independence, between the two groups over time. Compliance rates with the iMHere system and differences in phone usage did not significantly impact psychosocial outcomes.  While the iMHere system demonstrated effectiveness in reducing UTIs, it did not significantly influence other health or psychosocial outcomes. | Medical management |
| de Laat et al. [79]  2017 | To describe the extent of health activation and self-management behavior in paraplegics to prevent pressure ulcer's and associations between this behavior and patient characteristics. | Higher levels of education and the degree of paraplegia were positively associated with health activation, with odds ratios of 2.2 (p = 0.017) and 2.8 (p = 0.036), respectively. However, patients with a history of PUs did not show significant differences in health activation compared to those without a PU history. A discrepancy was observed between patients' intended and actual behaviors to prevent PUs.  Statements related to self-management behaviors revealed statistically significant differences between patients with and without a PU history for specific behaviors, such as maintaining correct positioning (p < 0.01), routinely checking the skin (p < 0.05), and using pressure-relieving devices (p < 0.001). Positive correlations were found between higher PAM stages (health activation levels) and the number of self-management behaviors reported (r = 0.152, p = 0.026), as well as between PAM stages and patients' opinions about health (r = 0.179, p = 0.024) and education (r = 0.192, p = 0.014).  These findings highlight the influence of education and injury level on health activation and underscore the gap between intended and actual self-management behaviors. | Medical management |
| Liu et al. [84]  2023 | To assess the effect of a self-management intervention delivered by mobile application (APP) for depression among community-dwelling  individuals with spinal cord injury (SCI). | The intervention group received five one-on-one sessions at weeks 2, 4, 6, 8, and 12, which included online assessments, health education, interdisciplinary referrals, and interactive support. Follow-up nurses provided guidance and referrals as needed. The control group received usual care and a single telephone follow-up at week 12, focusing on skin care, defecation management, self-care, and function training. Depression was measured using the Beck Depression Inventory-II (BDI-II) at weeks 12 and 24 post-discharge, with higher scores indicating greater depression. Baseline data, including demographic and injury-related information, were also collected. Of the 98 participants (49 in each group), the majority were male (82.7%), married (83.7%), and unemployed (94.9%), with an average age of 41.71 years. Trauma was the primary cause of injury (90.8%), and nearly half had complete spinal cord injuries (49.0%). The thoracic region was the most common injury site (46.9%). The intervention group showed a significant reduction in depression scores, with a decrease of 5.76 points at week 24 compared to the control group (95% CI = -9.97, -1.54; P = .007). Effect sizes indicated a small effect at week 12 and a moderate effect at week 24. These findings suggest that the nurse-led multidisciplinary intervention delivered via the "Together" app effectively reduced depression levels in individuals with SCI, highlighting its potential as a valuable tool for post-discharge mental health support. | Medical management  Emotional management |
| MacGillivray et al. [56]  2020 | To determine the feasibility of implementing and evaluating a self-management mobile app for spinal cord injury during inpatient rehabilitation and following community discharge. | The study assessed feasibility through recruitment rates, retention, intervention adherence, physical usability, adverse events, and the administration of outcome measures. The recruitment rate and retention met the study criteria, with over 80% of participants retained from admission to discharge and from discharge to 3 months post-discharge. Intervention adherence, measured by the number of times participants saved data in the app, varied between inpatient rehabilitation and community discharge, with a decrease in data entries after discharge. Physical usability of the app was evaluated in relation to participants' Spinal Cord Injury Independence Measure-III (SCIM-III) scores. While SCIM-III scores did not significantly correlate with app usage during inpatient rehabilitation, a positive trend was observed in the community. No adverse events related to the intervention were reported. Outcome measures, including the Hospital Anxiety and Depression Scale (HADS) and self-management confidence ratings, were administered effectively. Most participants scored within the 'normal' range on the HADS, though some reported scores indicating anxiety or depression. App usability varied, with some participants using the app independently and others requiring caregiver assistance or stylus use. Participants reported improved confidence in bowel management over time, with trends of improvement in bladder management, activities of daily living (ADL), and pain management. Medication management was rated as the most important area for addressing self-management. | Medical management |
| Meade et al. [68]  2016 | To examine the feasibility of administering an individual, in-person version of Health Mechanics, an innovative self-management program designed to teach individuals with spinal cord injury to maintain physical health and prevent secondary conditions. | Effectiveness was measured using the Spinal Cord Injury Secondary Conditions Scale (SCI-SCS), Patient Health Questionnaire (PHQ-9), Social Problem-Solving Inventory – Revised: Short (SPSI-R:S ), Disability Management Self-Efficacy Scale (DMSES), and a knowledge measure about managing SCI and preventing secondary conditions. A process evaluation gathered participant feedback on the study's assessment procedures and intervention components. Participants who completed the evaluation provided positive feedback, reporting that they learned and applied at least one skill from the program and believed they benefited from participating. Many noted that the program would be particularly beneficial for individuals earlier in their experience of living with SCI. Feasibility was assessed by examining participant recruitment, retention, and completion rates, as well as the mode of intervention delivery. | Medical management |
| Mortenson et al. [57]  2019 | To describe stakeholder perspectives on the development of a functional mobile app to facilitate self-management skills needed to prevent secondary complications following recent spinal cord injury during inpatient rehabilitation. | Three main themes emerged from the feedback: Being individualized and user-Friendly: Stakeholders emphasized the need for the app to be simple, accessible, and user-friendly to ensure broad adoption. They highlighted the importance of flexibility, allowing tools to be tailored to individual needs. Suggestions included simplifying and merging tools to reduce clutter while maintaining a variety of features to address diverse self-management needs. The goal was to make the app accessible to users with varying levels of health literacy. Targeting goals to promote self-management: Goal setting was identified as a critical component of self-management. Stakeholders recommended that the app encourage users to define their own goals based on their health needs and confidence levels. Tools for goal-setting, progress tracking, and motivational features, such as quotes and confidence tracking, were seen as essential for fostering independence and empowering users to take control of their health outcomes. Improving participation and gaining support to facilitate lifestyle change: Stakeholders stressed the importance of promoting community participation and social support through the app. They suggested features that encourage leisurely pursuits, balance work, school, family life, and recreational activities, and provide options tailored to individual needs. Social support tools, such as tracking social contacts and fostering community engagement, were seen as vital for addressing social isolation post-discharge and improving long-term health outcomes. | Medical management |
| Munce et al. [50]  2016 | To understand the meaning of self-management in traumatic spinal cord from the perspectives of individuals with traumatic spinal cord and their caregivers as well as acute care/trauma and rehabilitation managers. | Two overarching themes emerged: internal responsibility attribution and external responsibility attribution.  Internal responsibility attribution: Participants emphasized the importance of wellness awareness, including lifestyle practices such as good nutrition, exercise, and relaxation, as essential for maintaining health after SCI. Self-management also involved actively monitoring for secondary complications, such as skin issues, and taking preventive measures. The tension between striving for independence and the risk of injury was highlighted, with individuals balancing autonomy and safety. Managers noted the role of directing caregivers, often spouses, in providing care, emphasizing effective communication and guidance. Across all groups, self-management was viewed as taking ownership of one's health, including scheduling medical appointments and seeking necessary treatments.  External responsibility attribution: Managers linked self-management in SCI to established chronic disease self-management programs, such as the Stanford Chronic Disease Self-Management Program, which they saw as applicable to SCI. Participants also recognized the critical role of caregivers, who needed a diverse skill set to assist with daily living activities and help prevent, monitor, and manage secondary complications. | Medical management  Role management |
| Munce et al. [59]  2017 | To determine the implementation considerations for a targeted self-management program for individuals with spinal cord injury from the perspective of a national stakeholder advisory group using the Theoretical Domains Framework (TDF) as a guide. | The findings were organized into several domains, reflecting the diverse factors that influence the design and delivery of such a program. 1) Knowledge Domain: Participants stressed the importance of providing accurate, timely, and accessible information tailored to the readiness and health literacy levels of individuals with SCI. Information should be presented clearly, avoiding jargon, to ensure understanding. 2) Skills Domain: The need for certification and training for program facilitators, including peer mentors, caregivers, and healthcare professionals, was highlighted. The program should accommodate participants with varying skill levels and age ranges. 3) Social/Professional Role and Identity Domain: Establishing connections with primary care providers was seen as crucial to ensure awareness of the program and facilitate referrals. 4) Beliefs About Capabilities Domain: Peer support within the SCI community and the use of social media were identified as valuable tools for promoting the program and encouraging adoption. The program's labeling and messaging should address potential unfamiliarity with self-management concepts. 5) Beliefs About Consequences Domain: The program's goals and outcomes should align with the expectations of individuals with SCI, with input from this group shaping the program's content and objectives. 6) Reinforcement, Intentions, and Goals Domains: Considerations included tailoring the program to the stage of injury (newly injured vs. long-term), assessing readiness for self-management, and customizing the program to meet individual needs. 7) Memory, Attention, and Decision Processes Domain: The program should provide easily accessible content and fit into the schedules of individuals with SCI to ensure usability and engagement. 8) Environmental Context and Resources Domain: Leveraging existing resources, such as peer support networks, and addressing governance and ownership of the program were emphasized. 9) Social Influences Domain: Privacy and security measures for online delivery were highlighted, along with the potential for misinformation. The influence of caregivers and the need for their own self-management support were also discussed. 10) Optimism/Emotion Domain: The program should address sensitive topics like bladder management and depression while considering potential embarrassment or stigma, particularly among younger participants. 11) Behavioral Regulation Domain: Key content areas included stress management, secondary conditions, pain, coping strategies, locus of control, social roles, problem-solving, and action planning. Online tools were seen as valuable for engaging individuals in their self-management. | Medical management |
| Munce et al. [58]  2014 | To determine the importance attributed to the components of a self-management program by individuals with traumatic spinal cord injury and explore their views/opinions about the delivery of such a program. | Participants rated several components as "very important," including exercise (53.5%), nutrition (51.5%), pain management (44.4%), information/education on aging with SCI (42.4%), communicating with healthcare professionals (40.4%), problem-solving (40.4%), transitioning from rehabilitation to the community (40.4%), and confidence in reducing secondary complications/promoting wellness (40.4%). Overall, 74.7% of participants considered the development of a self-management program for traumatic SCI as "very important" or "important."  Participants expressed clear preferences for program delivery. The majority (39.4%) favored an internet-based format, with 29.3% of these preferring a one-on-one approach. Regarding program construction, 74.7% believed grouping individuals with similar injury levels would be most effective, while 40.4% emphasized the importance of similar age groups. Timing for introducing the program varied, with 42.4% favoring the rehabilitation period and 29.3% preferring the transition from rehabilitation to the community. Follow-up was deemed essential by 88.9% of participants, with 31.3% preferring regular contact with a healthcare professional. For program leadership, 38.4% suggested a combination of healthcare professionals and individuals with SCI, while 41% indicated that organizations like SCI Canada should organize the program.  Variations based on time since injury were noted, with individuals further post-injury more likely to rate modules on aging with SCI and relationship issues as important. | Medical management |
| Munce et al. [49]  2014 | To understand the perceived facilitators and barriers to self-management to prevent secondary complications. | The findings highlighted key factors that either supported or hindered effective self-management.  Facilitators to self-management: Participants emphasized the importance of physical and emotional support from caregivers, who assist with daily activities, secondary complication prevention, and emotional encouragement. Peer support programs, such as those offered by SCI Canada, were seen as highly beneficial, particularly when pairing newly injured individuals with experienced peers who share similar characteristics like age, sex, and injury level. A positive outlook and acceptance of the injury were also identified as critical facilitators, with work or volunteering contributing to maintaining this mindset. Additionally, maintaining independence and control over care, such as through access to transportation, was seen as essential for effective self-management.  Barriers to self-management: Caregiver burnout emerged as a significant barrier, with family members often taking on dual roles as spouses and caregivers without adequate support. Funding and policy limitations, such as insufficient funding for homecare services and disparities based on injury mechanism, were also identified as major obstacles. Accessibility issues, including difficulties accessing buildings and medical facilities, further hindered self-management. Physical limitations and secondary complications inherent to SCI were noted as barriers, as were challenges in achieving a positive outlook or mood, which could be exacerbated by traumatic brain injuries or changes in personality and motivation. | Medical management  Emotional management |
| Newman et al. [69]  2019 | To develop educational content and pilot test the use of tablet computers (iPads), online content management platform (iTunes U) and video conferencing (FaceTime) for delivery of a peer supported, spinal cord injury self-management intervention, using a community-engaged research approach. | The study evaluated the usability and acceptability of these tools. Participants with impaired hand function required adaptive equipment, such as styluses, to effectively use the iPad, but overall found it accessible and user-friendly. Suggestions for improvement included adding an introductory navigation video. The iTunes U platform and course content received favorable ratings for usability and acceptability, though course attractiveness was rated less favorably. Participants appreciated the engaging, humorous, and relatable videos but recommended increasing font size and featuring more people in the videos. For video chat (FaceTime), participants with impaired hand function faced challenges holding the iPad at their preferred viewing angle, indicating a need for additional equipment. Connectivity issues in rural areas prevented one participant from engaging, but once connected, FaceTime worked well, and participants had positive perceptions of the experience. | Medical management |
| Oh et al. [70]  2023 | To examine how digital  technology could improve SCI population’s adherence to pressure relief (PR) exercises. | Three key themes emerged from the findings: 1) Unique experiences to similar problems: Participants had diverse experiences with pressure injuries (PIs), varying in causes and severity. Understanding the severity and life-altering consequences of PIs motivated some to actively engage in PR exercises. Participants relied on body awareness and adapted their routines to fit their post-injury lifestyles, highlighting the importance of personalized approaches. 2) Increasing PR adherence through a personalized experience: Participants responded positively to reminders, which encouraged higher adherence to PR exercises. SMS reminders were preferred due to their simplicity and accessibility. Personalized reminder schedules and visual feedback were particularly effective, with participants noting that reminders helped them stay consistent, even during busy periods. 3) Gamifying and visualizing PR adherence: Gamification features, such as rewards and positive reinforcement, increased engagement and adherence. Some participants used emojis and visual elements for self-reporting, adding a sense of personal motivation and entertainment to the process. | Medical management |
| Olney et al. [71]  2019 | This paper reports the iterative redesign, feasibility and usability of the Comprehensive Mobile Assessment of Pressure (CMAP) system's mobile app used by veterans with spinal cord injury. | Interview results: In Phase 1a (formative focus groups), five key themes emerged: (1) Veterans emphasized the importance of skin ulcer prevention and expressed interest in using the app for pressure relief and reminders. (2) Users wanted the app to be simple, especially for those with limited smartphone experience. (3) Integration and reliability were critical, with users desiring a more seamless system. (4) Real-time pressure mapping was seen as the app's primary asset. (5) Veterans viewed CMAP as a medical tool introduced by healthcare providers and had no major privacy concerns. In Phase 1b (usability study), participants with prior smartphone experience found the app more user-friendly, and most considered it intuitive and straightforward. In Phase 2 (field use feasibility interviews), participants highlighted the importance of skin ulcer prevention and monitoring tools. They valued features like live pressure mapping and weight-shifting reminders but noted inconsistent functionality due to interconnected components (e.g., wireless, Bluetooth, wires, batteries). Participants also suggested expanding the app's use to other high-risk surfaces and provided recommendations for disseminating the technology.  Survey results: System Usability Scale (SUS) scores ranged from 47.5 to 100, with a mean of 72.1, indicating varying usability levels. Challenges with technology consistency affected some scores. The User Experience Questionnaire (UEQ) scores were generally positive, though the "Dependability" scale received the lowest rating due to predictability concerns. Skin checks using the pressure-sensing mat showed no changes in skin integrity. | Medical management |
| Pancer et al. [60]  2019 | To identify the preferred features of a Web-based self-management physical activity portal through stakeholder engagement with individuals with a spinal cord injury and health care professionals. | The findings were organized into behavior change techniques and modes of delivery.  Behavior change techniques: Participants emphasized the need for knowledge resources, including guidance on physical activity (e.g., home-based exercise tutorials, safety suggestions, and evidence-based guidelines) and strategies to overcome barriers like cost, equipment availability, and physical accessibility. They also highlighted the importance of understanding the risks and benefits of physical activity, with examples of individuals with SCI successfully engaging in physical activity (e.g., photos, testimonials, and mentors) serving as inspiring models. For self-regulation strategies, participants preferred tools for action planning (organizing when, where, and how to incorporate physical activity), goal setting (specific, meaningful, and realistic objectives), and tracking progress to measure improvements and stay accountable. Opinions on rewards were mixed, with some appreciating extrinsic motivators, while others felt intrinsic health benefits were sufficient. Similarly, reminders were seen as encouraging by some but stressful by others.  Modes of delivery: Participants valued interactivity, including peer platforms like discussion forums or chat groups for sharing resources and fostering a sense of belonging, as well as access to healthcare professionals via phone, email, or Skype for reassurance and continuity of care. Regarding format, participants preferred visually appealing content, such as photos, videos, colorful diagrams, and succinct text, as most identified as visual learners. They also emphasized the importance of clear, positive language and the availability of resources in French. A user-friendly website with structured, easily accessible content was seen as essential for efficient navigation. | Medical management  Role management |
| Pilusa et al. [87]  2021 | To explore how people with spinal cord injury prevent and manage secondary health conditions. | Thematic content analysis revealed three main themes: 1) Prevention of secondary health conditions: Participants employed self-management strategies to prevent conditions such as pressure ulcers and urinary tract infections. These strategies included pressure relief techniques, staying hydrated, and maintaining a healthy diet. Additionally, some participants relied on assistive devices, such as high-quality wheelchair cushions, to reduce the risk of developing pressure ulcers. These devices were seen as essential tools for prevention. 2) Management of secondary health conditions: Pain was identified as a common secondary health condition requiring active management. Participants used various approaches to cope with pain, including resilience, medication, and therapeutic interventions like hyperbaric therapy. These strategies aimed to alleviate pain and improve overall quality of life. 3) Challenging experience: Participants expressed frustration with the difficulty of preventing and managing secondary health conditions, particularly when some strategies proved ineffective. This frustration sometimes led to feelings of hopelessness, highlighting the emotional and psychological challenges associated with managing long-term health issues after SCI. | Medical management |
| Potiart et al. [83]  2020 | To develop and evaluate effectiveness of a smartphone application to assist the self-management of intermittent urinary catheter users | Feasibility: Participants expressed satisfaction with the app in the 1-month feasibility survey, finding it useful and demonstrating good adherence. Most continued to use the app after three months, with 48% finding the FAQ chatbot useful and 51.4% favoring its retention. User opinions remained consistent between the first and third months, with no significant differences.  Usability: Initially, most participants found the app easy to use and could access its various features. Half considered the app interesting, though only 40% reported improved bladder control with its advice. By the third month, more participants found the app difficult to use, and bladder control remained a challenge for many.  Catheter-related pain: Baseline pain scores were low (1.66 ± 1.8) and increased slightly to 1.8 ± 2.1 at one month and 2 ± 2.2 at three months. However, these changes were not statistically significant.  Leakage: Approximately 74% of participants reported rarely experiencing leakage, with no significant changes in leakage frequency observed between baseline, one month, and three months.  Urinary tract infection (UTI): About 30% of participants had experienced UTIs before joining the study. The number of participants reporting UTIs remained unchanged during the study, with no significant differences at the one-month and three-month surveys. | Medical management |
| Pryor et al. [88]  2021 | To describe the usual bowel care regimes of people living in the community with spinal cord injury and factors important for integrating bowel care into everyday life. | Participants had diverse bowel care routines influenced by their injury levels and types. Common practices included per rectum (PR) examination, enemas, digital stimulation, and manual evacuation if needed. Ten participants used aperients and enemas, while one was independent without aids. Some incorporated abdominal massage into their routines. Six participants received assistance from carers, while four managed independently. Bowel care frequency varied, with four participants performing it daily and seven every second day. Most combined bowel care with personal hygiene, typically in the morning, with the process taking 30 minutes to 3 hours. Four key factors were identified for successfully integrating bowel care into everyday life:1) Acceptance, motivation, and willingness: Participants emphasized the importance of accepting their situation, staying motivated to avoid complications, and taking responsibility for self-managing bowel care. Over time, they developed the necessary knowledge and skills. 2) Discipline: Establishing and maintaining a consistent routine, including performing bowel care at the same time and frequency, was crucial. Allowing sufficient time for the process was also important. 3) Proactive self-management: Participants proactively adapted their routines to solve problems, reduce complications, and accommodate changes in work schedules, diet, and age-related bowel function. 4) Fostering collaboration with carers: For those requiring assistance, collaborating effectively with carers was essential. Participants valued reliable, high-quality carers and emphasized the importance of training and problem-solving together. | Medical management  Role management |
| Raghavan et al. [90]  2003 | To estimate the point prevalence of pressure sores in a community sample of spinal cord injured patients who were followed up by a spinal injuries unit and to evaluate whether self-management strategies were associated with decreased risk of pressure sores. | The point prevalence of pressure sores was 23%, with 44% of affected individuals having more than one sore.  Factors associated with pressure sores: Logistic regression analysis identified current smoking and co-morbidity as factors independently associated with an increased risk of pressure sores. In contrast, regular skin inspection was linked to a decreased risk. Daily skin inspection, while associated with more stage I pressure sores, did not significantly affect the prevalence of stage II or higher sores. Lifting body weight while seated at least every hour was not associated with a reduced prevalence of pressure sores. Other factors, such as employment status, age, gender, neurological level, urinary incontinence, and fecal incontinence, were not significant predictors. Pressure sore staging: Among participants with pressure sores, 44% had multiple sores, and 82% of sores were located around the pelvic area (gluteal, sacral, or trochanteric). Additionally, 55% of respondents reported having at least one pressure sore of stage II or higher since their original injury. Pressure sore prevention strategies: Many participants reported using pressure-relieving cushions and mattresses to prevent pressure sores. Concurrent medical problems, such as ischemic heart disease, hypertension, and diabetes mellitus, were also noted among some participants. | Medical management |
| Shirai et al. [61]  2022 | To explore the experiences of individuals living with spinal cord injury/disease on the use of Pressure Ulcer Target (PUT), a mobile educational app for pressure injure prevention and management. | Strengths and weaknesses of PUT: Participants found the content highly beneficial and informative, aligning with information provided by healthcare professionals. However, some noted that the medical terminology was too scientific, potentially reducing user interest, and that certain sections were redundant. Navigation was a mixed experience; some found the layout logical, while others felt the module order was unclear and navigation cumbersome, especially for those with limited hand mobility. Aesthetically, participants appreciated the animations and diagrams but suggested more visuals in text-heavy sections and adjustable font sizes for older users. Target population for PUT: Participants had varied experiences with pressure injuries (PIs), ranging from severe cases to no prior experience. They emphasized the need to educate healthcare providers and support workers about PIs. Opinions on the ideal timing for introducing PUT differed, with some advocating for early introduction during inpatient rehabilitation and others suggesting it be introduced after the initial emotional impact of the SCI/D diagnosis had subsided. Key concepts and messages as motivators for using PUT: The concept of prevention was a strong motivator, with users appreciating information on prevention strategies and the impact of PIs on quality of life. Learning about the consequences of PIs on health and daily life encouraged engagement with the app. Recommendations for improvement: Participants suggested expanding content to include topics like physical activity, nutrition, behavioral changes, self-cleaning practices, and equipment modifications. They recommended enhancing visuals with real images, more animations, and video content to balance text and improve engagement. Accessibility improvements included making the "Check" List printable, adding audio features, providing a zoom-in function, and improving navigation guidance. | Medical management |
| Singh et al. [62]  2020 | The objective of this study was to explore patients' perspectives on the usability of this self-management app. | Participants highlighted three key aspects of usability: accessibility, intuitive navigation, and flexibility. Accessibility: Participants emphasized the importance of making the app accessible to users, noting that technological knowledge significantly influenced usability. Ensuring the app was easy to use for individuals with varying levels of tech-savviness was a priority. Intuitive navigation: Participants stressed the need for an intuitive interface, including a simple tool selection menu. Multitasking capabilities were important, and they recommended incorporating alerts and reminders to enhance functionality and user engagement. Flexibility: Flexibility in data entry, the ability to backtrack and update information, and offline functionality were identified as essential features. These elements allowed users to interact with the app in a way that suited their individual needs and circumstances. System Usability Scores (SUS): The mean SUS score at discharge was 78.1 (SD: 15.6; Range: 45-100), indicating good usability. At three months post-discharge, the mean SUS score was 71.6 (SD: 12.7; Range: 55-92.5), showing a slight decline but still reflecting acceptable usability. The difference in SUS scores between the two time points was not statistically significant (p = 0.219), suggesting consistent usability over time. | Medical management |
| Skeels et al. [39]  2017 | To describe the roles fulfilled by peer health coaches (PHCs) with spinal cord injury during a randomized controlled trial research study called 'My Care My Call', a novel telephone-based, peer-led self-management intervention for adults with chronic spinal cord injury 1+ years after injury. | PHCs fulfilled three primary roles—Role Model, Supporter, and Advisor—using specific communication tools (CTs) and information delivery strategies (IDSs). Role model: PHCs used reflective listening (RL) and shared stories (SS) to empathize and connect with peers over shared SCI experiences. They shared personal stories and opinions, modeling effective self-management and healthcare communication skills. This role helped build trust and provided relatable examples for peers to follow. Supporter: In this role, PHCs employed RL, SS, in-between call support, affirmations, and relationship-building IDSs to foster trust and confidence in peers' self-management abilities. They encouraged peers, identified supportive friends and family, and provided text messaging support between calls to maintain engagement. Advisor: As Advisors, PHCs used RL, SS, action planning (BAP), resource reviews, and in-between call text reminders. They sent tailored Postcall Support Packages and focused on teaching and strategizing with peers to enhance self-management skills. PHCs shared knowledge about health, SCI, assistive technology, and healthcare navigation, while also sending goal and action plan reminders via text. PHC role patterns over time:  The study examined how PHC roles evolved over the 6-month intervention: Months 1-2 (Weekly Calls): PHCs primarily acted as Supporters, followed by Advisors and Role Models. Months 3-4 (Bi-weekly Calls): The Supporter role remained dominant, with Role Model and Advisor interactions continuing. Months 5-6 (Monthly/Bi-weekly Calls): PHCs reported the highest number of interactions as Supporters and Advisors, with Role Model interactions declining. | Medical management  Role management |
| Starosta et al. [74]  2024 | To understand how individuals engaged with peers in the context of a self-management program for SCI. | Resource sharing: Participants exchanged valuable resources related to SCI, such as websites, equipment, exercise routines, adaptive sports, mindfulness practices, and caregiving tips. This peer-to-peer information exchange often included expressions of care and validation of shared experiences. Storytelling played a significant role, with participants sharing how achieving personal goals led to health improvements, creating a sense of connection and motivation. Skill building: The SCI Thrive course guided participants in setting and achieving goals related to managing life with SCI and improving quality of life. Participants engaged in mindfulness practices, such as guided imagery, breathing exercises, and autogenic relaxation. The group setting provided accountability, encouraging more frequent engagement with these practices and fostering skill development. Problem solving: Participants shared solutions to common challenges faced by individuals with SCI, leveraging collective wisdom to address issues. This theme often overlapped with emotional support, as participants expressed empathy and encouragement while discussing practical solutions. Bearing witness: This deeper, process-level theme involved participants emotionally connecting with others who shared similar SCI-related challenges. The term "bearing witness" described the act of sharing and responding to experiences that individuals without SCI might not fully understand. Participants frequently noted that the group was the first time they had engaged with a community that truly understood their lived experiences, highlighting the importance of emotional connection and shared understanding. | Medical management  Role management |
| Van Gaal et al. [81]  2022 | To explore how individuals with spinal cord injury self-manage the prevention and treatment of pressure ulcers and to provide insight into experiences with self-management support | The findings were organized into three main areas: managing medical tasks, managing emotional impacts, and integrating prevention and treatment into social and everyday life. Managing medical tasks: Participants emphasized daily preventive measures, such as skin inspection and pressure relief, to avoid pressure ulcers. However, they often relied on assistance from informal caregivers, as healthcare professionals in the community sometimes lacked sufficient knowledge about SCI-specific prevention. Rehabilitation courses and a healthy lifestyle, including exercise and diet, were seen as important preventive strategies. Support aids like seating cushions and special mattresses were commonly used, though not always consistently. Peers played a key role in sharing information about technological aids. Treatment experiences varied, with some requiring extensive hospitalization and surgeries, while others were treated at home. Participants stressed the importance of being actively involved in treatment decisions and tailoring interventions to their quality of life goals. Vigilance in identifying high-risk situations, such as medication changes or improper seating, was crucial to prevent recurrence. Managing emotional impact: Participants often felt dependent on others for tasks like skin inspection and dressing, which could lead to stress and anxiety, especially when disagreements arose with caregivers. Long hospital stays for pressure ulcer treatment were described as lonely and emotionally challenging, highlighting the need for emotional support from peers and healthcare professionals. Honesty and emotional support from nurses were highly valued, and peers were seen as essential for providing perspective and coping strategies. Managing prevention and treatment in social and everyday life: Balancing pressure ulcer prevention and treatment with family life required coordination with caregivers and care organizations. Participants highlighted the challenges of maintaining family connections during hospitalization. At work, preventive measures like changing positions were difficult to implement due to busy schedules, and participants often felt pressured to continue working despite pressure ulcers. Engaging in hobbies or sports also required careful management of materials and positions to avoid exacerbating pressure ulcers. | Medical management |
| Wang et al. [85]  2023 | To evaluate the skin self-management of community-dwelling patients with spinal cord injury and to explore the related independent influencing factors. | The study included 110 participants, predominantly male (81.8%) and under 60 years old (89.1%). Most had primary or secondary education (70.9%), and 88.2% had SCI due to trauma. The median time since injury was 15 months, with the majority having paraplegia (81.8%). Comorbidities like hypertension, diabetes, and chronic nephritis were present in 15.5% of patients, and 11.8% had experienced pressure injury recurrence. Revised SMnac scores: Lower scores indicated poorer skin self-management. The mean Revised SMnac score was 48.74 ± 15.91, with subcategory scores as follows: skin check (8.60 ± 4.25), preventing pressure ulcers (28.42 ± 9.89), and preventing wounds (11.72 ± 3.77). Factors influencing self-management: Univariate analysis revealed worse self-management in patients aged 60–90 years, those with a medical expense reimbursement ratio of 30%–80%, and those with comorbidities. Correlation analysis showed significant relationships between self-management and functional independence, knowledge about skin self-management, attitude toward skin self-management, and self-efficacy. The strongest correlation was between preventing pressure ulcers and knowledge about skin self-management.  Linear regression analysis identified key predictors of skin self-management: Higher levels of knowledge about skin self-management (β = 0.281) were associated with better self-management. A higher medical expense reimbursement ratio (>80%) (β = -0.211) was linked to improved self-management. Greater self-efficacy (β = 0.178) also positively influenced self-management practices. | Medical management |
| Widerström et al. [73]  2023 | To develop and evaluate a pain education resource (SeePain) for individuals with SCI and their significant others | Stakeholder feedback was incorporated to improve the resource's content, comprehensibility, and format.  Content of the SeePain resource: Participants suggested adding more relevant resources, such as a list of healthcare providers specializing in chronic pain management for SCI. They also recommended including information on pain triggers, self-management strategies, and support for significant others, who often bear emotional burdens. Additional requests included more details on the use of cannabis and opioids for pain management, a glossary of medical terms with definitions and phonetics, and real-life examples of pain experiences to make the content more relatable and practical. Comprehensibility of the SeePain resource: Feedback emphasized the need to simplify complex medical jargon and use plain language to ensure the resource is accessible to all users. Visual aids, such as diagrams, pictures, and clear explanations, were recommended to enhance understanding and engagement. Format of the SeePain resource: Participants suggested creating both comprehensive and shorter versions of the resource to suit different settings, such as early inpatient care or quick doctor visits. Adjustments to layout and design, including color choices, picture selections, and section organization, were made to improve user-friendliness. Additional formats, such as short videos, PowerPoint presentations, and smartphone apps, were proposed to make the information more accessible and engaging. Revised SeePain resource: Based on stakeholder feedback, the SeePain resource was revised into two modules: Module 1 (23 pages): Focused on basic pain education, covering the nature of pain, types of pain, and real-life examples. Module 2 (43 pages): Provided detailed content on pain management strategies, including non-pharmacological approaches, evidence-based treatments, and information on cannabinoids and opioids. It also included a glossary of terms and multiple resources for further support. | Medical management |
| Wilde et al. [40]  2011 | To identify and describe issues of intermittent urinary catheter users for future self-management research and/or training programs. | The findings were based on questionnaires and interviews, revealing key issues and experiences related to clean intermittent catheterization (CIC). Participants performed catheterizations an average of 5.6 times per day, with most using plastic uncoated catheters and some opting for latex, hydrophilic, or silicone catheters. Lubrication was used by 22 participants, while 12 did not use any. Over the past year, participants experienced an average of 2.3 urinary tract infections (UTIs), with three individuals hospitalized for a total of 20 days due to UTIs. Leakage was reported by 27 participants, with over a third experiencing it daily. Catheter-related pain was reported by 13 individuals, with no significant association between catheter type and pain.  Interview themes: Knowing the body: Participants emphasized the importance of understanding their bodies, including recognizing sensations that indicated the need for catheterization. They discussed monitoring fluid intake, urine color, UTI symptoms, and maintaining cleanliness. Practicing CIC: Participants highlighted the need for individualized approaches to CIC and the importance of practice in mastering the technique. They shared insights on catheterization positions, clothing considerations, and the challenges of adapting to CIC. Limited options in catheters and equipment: Participants described difficulties in finding suitable catheter equipment, with some actively seeking information about products and others facing limited choices. Costs and insurance coverage were also significant concerns. Inaccessible bathrooms: Many participants expressed frustration with the lack of accessible bathrooms in public and private spaces. Issues included limited space, lack of privacy, and cleanliness, leading some to catheterize in unconventional locations. Hassles: Participants discussed various hassles, including travel difficulties, adapting to social situations, and the impact on intimate relationships. Leakage during sex was a source of embarrassment for many. Adjustment in making CIC a part of life: Participants shared advice for those new to CIC, emphasizing the importance of acceptance and adjustment. They discussed how CIC became integrated into their lives and offered coping strategies for lifestyle changes. | Medical management |
| Wilde et al. [42]  2016 | The purpose of this study was to evaluate the feasibility of a new web-based intermittent catheter self-management intervention. | Participants demonstrated significant improvements in self-management of neurogenic bladder dysfunction, as well as increases in catheter-related self-efficacy and quality of life scores. However, there were no significant changes in the frequency of urinary tract infections (UTIs) or catheter-related pain. Feasibility, acceptability, and usability: The intervention components, except for the forum, were highly rated for usefulness, satisfaction, and usability. Participants valued the intake and output diary, journal, educational materials, and calls with the study nurse. Responses to the forum were mixed, with some participants finding it less engaging. Seven out of 22 respondents used mobile phone modifications, with mixed ratings of usefulness, though six of the seven mobile users described their experience positively. Most participants found the study length satisfactory, though some suggested it could have been longer. Participants appreciated working with the study nurse and the one-on-one interaction. Suggestions for improvement included visual conversations with the nurse and enhancements to the mobile application.  Usage: Website usage data showed that participants viewed all pages of the educational booklet, with the most frequently visited pages covering optimal fluid intake, catheterization intervals, and recognizing UTI symptoms. Over half of the total time spent on the website was devoted to using the urinary diary. Changes in self-care management: Participants reported positive changes in self-care management, including increased awareness of fluid intake, trials of new catheter products, greater confidence, and adjustments to catheterization frequency. Many participants had not tracked fluid intake and urine output since their initial rehabilitation and found the urinary diary valuable for self-management. Most participants tracked intake and output for at least three consecutive days, with entries typically made within 24 hours of the corresponding events. Forum: Eight participants actively engaged in the forum, which covered a wide range of topics related to catheterization and SCI self-management. While some participants suggested improvements, such as visual conversations with the nurse and mobile app modifications, most indicated they would recommend the program to others. | Medical management |
| Wilde et al. [41]  2015 | To report the development of a Web-based self-management intervention for intermittent urinary catheter users and pretesting with four adults with spinal cord injury living in the community. | The intervention website included several components designed to enhance self-management skills and improve outcomes.  Educational materials: The Educational Materials section featured a 23-page illustrated Educational Booklet, adapted from a previous self-management program for indwelling catheter users and modified based on literature and qualitative research. The booklet covered topics such as optimal fluid intake, catheter selection, urinary tract infection (UTI) prevention, social support, and sexual activity, providing comprehensive guidance on self-management. Personal data section: This interactive section allowed participants to set goals, record catheterization frequency, track intake and output (I&O), maintain a journal, and view summary tables and graphs of their data. It provided a detailed overview of their self-management efforts, helping them identify patterns and trends in fluid intake, urine output, activities, and catheterization frequency. Study nurse consultations: Participants had two phone consultations with a study nurse. The first consultation focused on teaching participants how to use the online diary for self-monitoring. The second consultation reviewed self-monitoring results, discussed I&O data, and suggested strategies to prevent problems. A third consultation at three months addressed additional issues and provided further guidance. Peer-Led online forum discussions: Once enough participants enrolled, online forums were made available to facilitate peer-led discussions on various self-management topics. Peer leaders guided the discussions, and clinical experts were available to answer complex questions, fostering a supportive community for shared learning and problem-solving. Mobile phone modification: Based on feedback from pretesting participants, the study team developed a mobile phone modification to make the intervention more accessible and user-friendly. This enhancement aimed to improve convenience and engagement for participants using mobile devices | Medical management |
| Zanini et al. [77]  2020 | This study aimed to identify styles of prevention that individuals with spinal cord injury adopt to deal with the risk of developing pressure injures. | Three main prevention styles were identified: Thoughtfuls, Selectives, and Delegators. Each style reflected different levels of engagement, knowledge, and attitudes toward prevention. Prevention styles: Thoughtfuls: These individuals were highly proactive and disciplined, adhering to all recommended preventive measures. They prioritized prevention over other activities, using assistive devices consistently and anticipating risks. Thoughtfuls demonstrated strong organizational skills and a commitment to staying healthy. Selectives: Selectives performed only a subset of preventive measures, balancing prevention with other life commitments. They relied on personal experience and intuition, sometimes deviating from healthcare professionals' recommendations. Selectives carefully observed their bodies and adapted their behavior to their strengths and weaknesses. Delegators: Delegators were relatively inactive in prevention, often delegating tasks to caregivers. They preferred passive measures like air-flow beds over active measures like skin inspections. Delegators had high expectations for assistive devices and were frustrated when PIs occurred despite their use. Knowledge and attitudes toward prevention: Extensive vs. basic knowledge: Thoughtfuls and Selectives had extensive knowledge of PIs and preventive measures, enabling them to recognize and respond to risks effectively. Delegators had limited knowledge and discussed only basic preventive measures. Personal responsibility vs. Delegation: Thoughtfuls and Selectives viewed prevention as a personal responsibility, while Delegators often blamed others, such as homecare services, for PI development. Priority of prevention: Thoughtfuls considered prevention a top priority, understanding the seriousness of PIs. Selectives valued prevention but aligned it with their life preferences. Delegators did not prioritize prevention as highly.  Perceptions of prevention: Participants had varying perceptions of prevention, with some viewing it as a habitual behavior and others emphasizing the effort and commitment required. Collaboration with healthcare professionals (HPs): Thoughtfuls and Selectives: These individuals actively sought support from HPs, building networks of experts for advice and care. They preferred HPs with SCI expertise and were proactive in seeking solutions. Delegators: Delegators favored more supervision from HPs, such as regular checkups or home visits, and were less proactive in seeking advice.  Attitudes toward life with SCI: Optimism: Thoughtfuls and Selectives were optimistic about their abilities and future, which motivated them to prioritize prevention. Self-Efficacy: Both groups expressed confidence in recognizing body signals and applying their knowledge. Thoughtfuls believed they could prevent PIs proactively, while Selectives felt they could react effectively to problems. Proactivity: Thoughtfuls were highly proactive, seeking peer input and optimizing preventive measures. Delegators tended to be more passive in both life and prevention. | Medical management  Emotional management |
| Zanini et al. [78]  2019 | To identify health professionals' perceived challenges in building and maintaining this partnership with patients, with a specific focus on how people with spinal cord injury and health professionals collaborate in the prevention and treatment of pressure injuries in spinal cord injury. | Three key challenges were identified: defining responsibilities and expectations, negotiating priorities and setting common goals, and building mutual trust and respect. Defining responsibilities and expectations: HPs emphasized that preventing PIs is primarily the responsibility of the individual with SCI, with their role being to empower, educate, and support self-management. Developing a sense of personal responsibility in individuals with SCI is crucial for effective prevention. However, HPs noted that this responsibility varies depending on factors like age or mental health status. Some individuals may lack personal responsibility due to trauma, psychosocial barriers, or a focus on self-determination over the responsibilities associated with SCI. In such cases, HPs might limit treatment options, opting for conservative measures due to the strictness of postoperative rehabilitation and limited surgical options. Negotiating priorities and setting common goals: Preventing and treating PIs often requires limiting an individual's freedom, such as enforcing bed rest or restricting travel. HPs sometimes need to convince individuals of the necessity of these limitations or negotiate solutions that balance medical necessity with the patient's priorities and quality of life. Setting personal, relevant objectives can motivate adherence to care plans. However, these negotiations can create tension, leading to frustration or a sense of helplessness for HPs. Building mutual trust and respect: Trust and a judgment-free relationship were deemed essential by HPs. Trust is critical because HPs rely on patient reports to verify adherence to recommendations in daily life. A trusting relationship encourages open discussions about sensitive issues related to prevention and treatment. HPs found that patients were more receptive to external support when suggested by someone with whom they had a strong personal relationship. Trust and respect are fostered through a good professional reputation, active listening, recognizing patient expertise, clear communication, confidentiality, and continuity of care. Dialogue and patience were considered more effective than issuing warnings in difficult situations. | Medical management |
| Zhou et al. [72]  2020 | To identify an approach that can be generally applied to improve the accessibility of mHealth apps. | Feedback and desired accessibility features: Participants provided detailed feedback on accessibility issues, including font size, spacing, button arrangement, color contrast, alternative data input methods, page navigation, handedness, and multimedia content. These insights guided the development of accessibility features tailored to the needs of individuals with disabilities. Accessibility features implemented: Based on participant feedback, the study team implemented several accessibility features in the iMHere 2.0 app. These included adjustable font sizes and styles, button and spacing adjustments, customizable color themes, and the option to select from provided answers instead of typing. Multimedia content was also added to enhance understanding and engagement. Performance with accessibility features: After implementing the accessibility features, participants found the app significantly easier to use. They required fewer attempts to complete tasks and reported improvements in clicking buttons, making selections, navigating the app, and understanding its content. These changes demonstrated the effectiveness of the accessibility enhancements. Usability: Participants rated the app's usability highly, with an average System Usability Scale (SUS) score of 90, indicating excellent usability. Positive feedback highlighted that improved accessibility directly correlated with better usability, making the app more user-friendly for individuals with disabilities. Other accessibility features: Participants with Bluetooth-capable power wheelchairs were able to navigate the app using their wheelchair controls, providing an alternative method for interacting with mobile devices. This feature further enhanced accessibility for users with limited mobility. | Medical management |
