## Supplemental Table 4 for "Trends in self-management research in spinal cord injury: A scoping review of study designs and findings"

| Themes | Subthemes | Contributing studies |
| --- | --- | --- |
| Individual factors | Knowledge at the cornerstone of SM | [36, 37, 39-41, 44, 45, 47-50, 54, 55, 57-61, 63, 66, 73, 74, 76, 77, 79, 81, 85, 86, 88] |
|  | Psychological well-being an important element in SM of SCI | [49, 74, 81, 86, 87, 89] |
|  | Self-management of SCI as part of daily life | [40, 45-48, 50, 54, 58-60, 68, 86, 88, 89] |
| Interpersonal and social influences | Patient-provider relationship influences SM | [47, 48, 77, 78] |
|  | Societal context shapes SM of SCI | [40, 45, 57, 59, 81, 86, 89] |
| Technological integration | Apps and programs enhance SM outcomes | [38, 42, 43, 56, 67, 75, 84] |
|  | Usability and feasibility elements influence the adoption of apps and programs | [36, 42, 43, 61, 62, 65, 69-72, 83] |
