## Supplemental Figure 1 for "Trends in self-management research in spinal cord injury: A scoping review of study designs and findings"

Cinahl

n = 950

Cochrane

n = 187

Science Direct	n= 204

Scopus

n = 1984

PubMed

n = 667

Identification

Duplicates

n = 1337

Screening

Records excluded

n = 2553

*Not about SM (n=940)*

*Before 2003 (n=571)*

*Not about SCI (n=384)*

*Not original study (n=282)*

*Not adult participants (n=110)*

*Book chapters (n=84)*

*Not found (n=50)*

*Protocol (n= 25)*

*Conference papers (n=25)*

*Not peer-reviewed (n=71)*

*Not English (n=5)*

*Editorial (n=4)*

*Pilot study (n=1)*

*Policy brief (n=1)*

Title and abstract screening

n = 2655

Eligibility

Records excluded

n = 50

*Not about SM (n=50)*

Full-text screening

n = 102

Included

Articles included in the synthesis

n = 52
