## Supplemental Figure 2 for "Trends in self-management research in spinal cord injury: A scoping review of study designs and findings"

**Theme**

**Code**

**Contributing Paragraph**

Significance of cultivating mutual trust

Providers' lack of knowledge about spinal cord injury and its medications affected their confidence in supporting medication management, while positive provider-patient relationships and trust facilitated better outcomes.

Providers' knowledge and confidence in managing SCI/D and related medications indirectly impacted adherence by shaping the quality of patient education and trust-building.

Patient-provider relationship influences SM

Trust is critical because providers rely on patient reports to verify adherence to recommendations in daily life. A trusting relationship encourages open discussions about sensitive issues related to prevention and treatment…Trust and respect are fostered through a good professional reputation, active listening, recognizing patient expertise…

Open communication and collaboration

Participants rated several components as "very important," including communicating with healthcare professionals (40.4%)

Follow-up was deemed essential by 88.9% of participants, with 31.3% preferring regular contact with a healthcare professional.

Health professionals emphasized that preventing pressure injuries is primarily the responsibility of the individual, with their role being to empower, educate, and support self-management

Definition of responsabilities and roles

Interprofessional collaboration, whether within multidisciplinary teams or through external consultations, was critical for effective medication management.
