## Supplementary figures and images for "Trends in self-management research in spinal cord injury: A scoping review of study designs and findings"

### Supplemental Figure 3

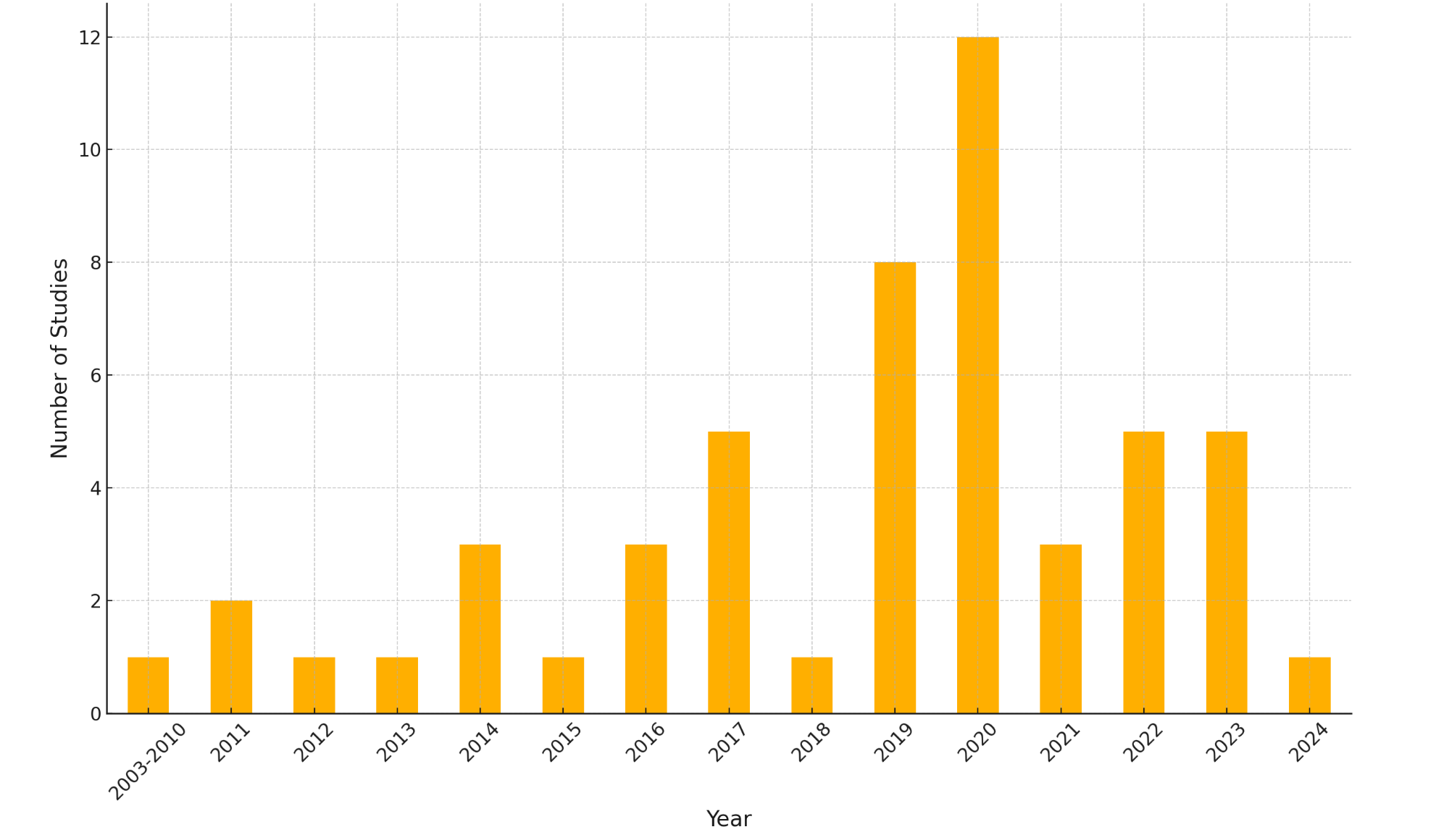

### Supplemental Figure 4

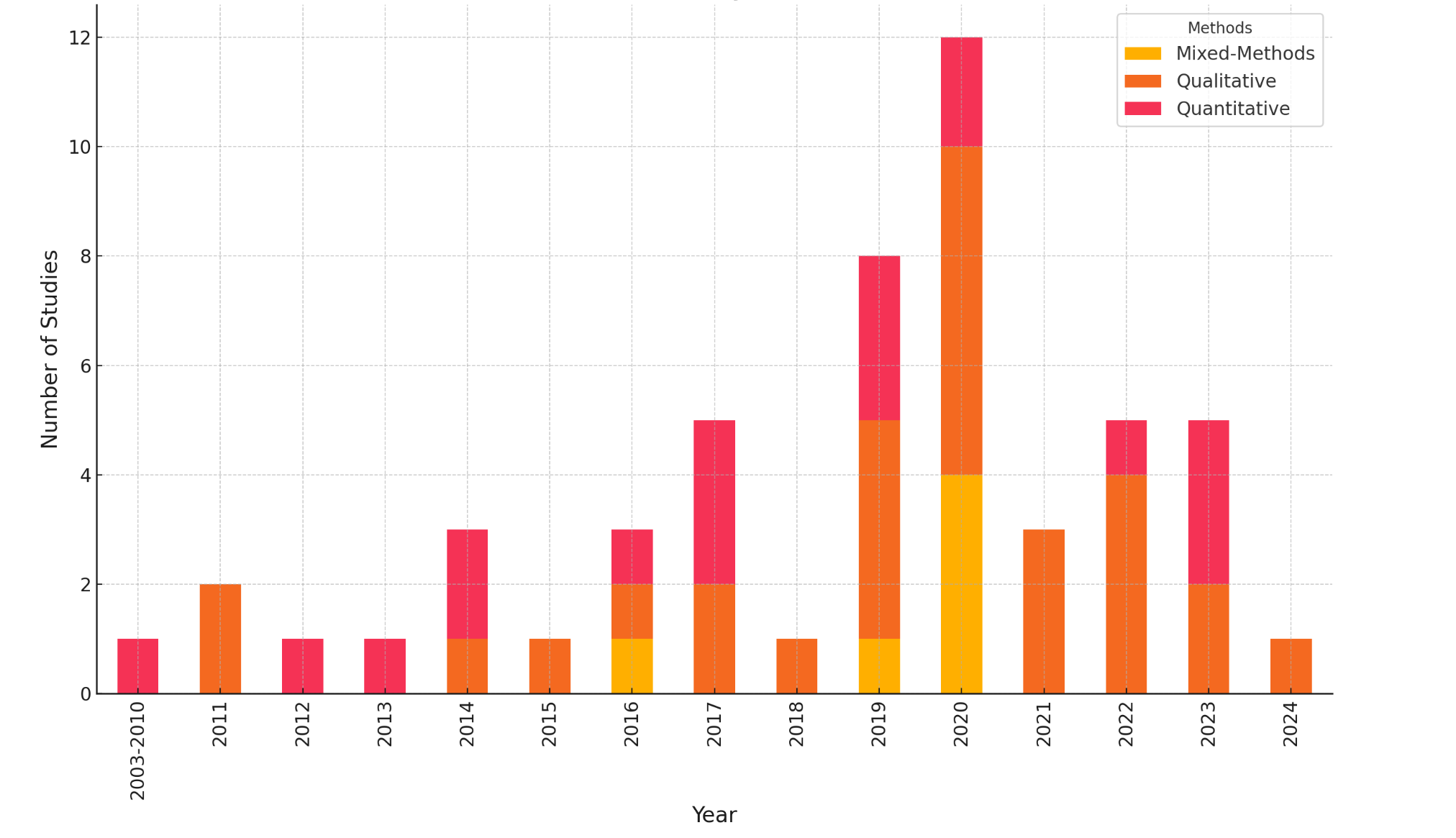
